# Clinical and genetic correlates of a circadian subtype of depression in the Australian Genetics of Depression Study

**DOI:** 10.64898/2026.02.23.26346917

**Authors:** Emiliana Tonini, Jacob J Crouse, Mirim Shin, Joanne S Carpenter, Brittany L Mitchell, Enda M Byrne, Penelope A Lind, Scott D Gordon, Richard Parker, Samuel J Hockey, Timothy To, Andrew Shim, Alexander Hill, Anna Treneman, Elizabeth M Scott, Jan Scott, Kathleen R Merikangas, Naomi R Wray, Nicholas G Martin, Sarah E Medland, Ian B. Hickie

## Abstract

**Background:** While commonly accepted depressive subtypes reflect phenotypic differences, there has been minimal progress in identifying discrete pathophysiological pathways, biomarkers or differential therapeutic approaches which effectively guide clinical management.

**Aims:** To test the biological validity and clinical utility of a circadian subtype of depression on the basis of clinical course, differential medication response (self-reported) and genetic risk profile.

**Methods:** Cross-sectional data were drawn from the nationwide, genetically-informative Australian Genetics of Depression Study. Participants were classified as having a “circadian” versus “non-circadian” subtype of depression on the basis of meeting criteria for at least three binary circadian features: social jetlag, seasonality, delayed sleep midpoint, evening chronotype, sleep inertia, and hypersomnia. Clinical course characteristics were compared. Associations with response to commonly prescribed antidepressants and polygenic risk scores (PGS) for mental disorders and sleep, circadian, metabolic and inflammatory traits, were investigated using logistic regression models.

**Results:** 2,604 participants (23%; 80% females; mean age=37.87±13.62) had a circadian subtype. These cases reported an earlier age of onset (*p*<0.001), more severe clinical features including hypo/manic-like and psychotic-like experiences, suicidality, psychological distress and somatic complaints (*p’s*<0.001), weight gain during depressive episodes (*p*<0.001), poorer response to SSRIs (OR=0.88 [0.82, 0.94]) and SNRIs (OR=0.89 [0.83, 0.97]) and more side-effects, compared to those with a non-circadian subtype. Having a circadian subtype was associated with higher PGS for attention-deficit/hyperactivity disorder (OR=1.11 [1.06, 1.17]), major depression (OR=1.11 [1.06, 1.16]), bipolar disorder (OR=1.09 [1.04, 1.14]), body mass index (OR=1.09 [1.05, 1.14]), triglycerides (OR=1.10 [1.06, 1.16]), interleukin-6 (OR=1.08 [1.03, 1.13]), higher insulin resistance (OR=1.08 [1.04, 1.13]), later sleep midpoint (OR=1.15 [1.10, 1.21]), insomnia (OR=1.08 [1.03, 1.13]), and later chronotype (OR=0.68 [0.65, 0.71]).

**Conclusion:** These findings support the face validity and potential clinical utility of circadian subtype of depression as a clinical profile. Pending independent replication, investigation of its biology and predictive utility are warranted.

## 1. Introduction

Major depressive disorder (MDD) is a heterogeneous condition with highly variable presentations and patterns of treatment response, likely reflecting multiple underlying pathophysiological mechanisms. Despite decades of research, most proposed clinical subtypes of mood disorders show only modest differences in prognosis or treatment response (1). Consequently, international treatment guidelines largely emphasise dimensions such as severity, persistence or recurrence (2, 3), often resulting in repeated trial-and-error prescribing of common antidepressants (4). Earlier detection of distinct illness pathways could enable more effective, personalised interventions at earlier clinical stages.

### Proposed circadian subtype of depression

Accumulating evidence implicates circadian disruption as a pathophysiological mechanism causing depressive symptomatology (5). Circadian disruption has been proposed as a marker for a distinct profile, termed “*circadian depression*” (6), which has garnered attention as a construct of interest (7). This cross-cutting profile is proposed to preferentially capture cases who have a circadian pathophysiology as a core contributor to their illness (6). Phenotypically, this circadian subtype is proposed to be characterised by disrupted 24-hour sleep-wake cycles, reduced daytime activity, low energy, and weight gain during episodes (6), and to be associated with early onset, bipolar-like features, poor response to common antidepressants (e.g., selective serotonin reuptake inhibitors [SSRIs]), and cardiometabolic/inflammatory comorbidities (8). Objective markers of circadian dysregulation have been reported in emerging mood disorders displaying circadian-like clinical features (9–11). Actigraphy-derived circadian measures are heritable (12), and genetic risk for bipolar disorder (BD) and MDD has been linked to delayed circadian rhythms (13). Circadian gene variants also appear to contribute to the development of mood disorders (14) and evening chronotype (a proposed feature of the circadian subtype) has been tied to poorer response to SSRIs and serotonin and norepinephrine reuptake inhibitors (SNRIs) (15).

### Current study

We leveraged a nationwide genetically-informative study of >10,000 adults with depression and aimed to (1) construct a circadian subtype phenotype using a selection of circadian-associated self-reported markers; (2) investigate the correlates of this circadian subtype including symptoms, clinical course, and self-reported treatment response to common antidepressants; and (3) examine associations between this circadian subtype and genetic risk for relevant mental disorders, physical health, and sleep/circadian traits. We hypothesised that the circadian subtype will be associated with more severe features (e.g., earlier age of onset), poorer response to conventional antidepressants, and higher genetic risk for bipolar disorder and metabolic and inflammatory markers, relative to a ‘non-circadian’ subtype of depression. Our overarching goal was to examine whether this construct could indeed be a valid and clinically useful phenotype to guide clinical future stratification efforts.

## 2. Methods

### a. Participants

Participants were from the Australian Genetics of Depression Study (AGDS), a nationwide cohort exploring genetic and psychosocial factors influencing the aetiology of depressive disorders and antidepressant response. The cohort profile is described elsewhere (16). Data from the first freeze (September 2016 to September 2018) were analysed. Ethical approval was granted by the Queensland Institute of Medical Research (QIMR) Berghofer Medical Research Institute Human Research Ethics Committee in Brisbane, Australia. Written informed consent was obtained.

A total of 20,689 participants (75% female; mean age=43±15 years [range: 18-90]) with a self-reported diagnosis of, or treatment for, a depressive disorder were recruited; 76% provided a saliva sample. Participants completed a survey comprising of a core module on depressive symptomatology and experiences with common antidepressants, and additional modules including one on sleep and circadian-related behaviours. The Composite International Diagnostic Interview Short Form was used to assess MDD according to Diagnostic and Statistical Manual of Mental Disorders (DSM)-5 criteria. The AGDS predominantly comprises adults with severe, recurrent depressive disorders, with only 4% reporting a single episode (16). Here, we only included participants of genetically-inferred European ancestry with complete data on PGS and on at least three circadian criteria (described below). A total of 11,380 participants (75% female; mean age=43.8±15.3 years) were included in this study (see Figure S1 for participant selection and Table S1 for comparison with excluded sample). Sensitivity analyses restricted to participants with complete data on all six circadian criteria (n=8,728) are presented in the Supplementary Materials (Figure S5 and Table S6).

### b. Circadian criteria

To identify participants with a circadian subtype, the following criteria were selected on the basis of availability, domain knowledge, and clinical expertise:

- <u>Seasonality</u>: Seasonality is proposed to be a marker of the circadian system’s sensitivity to photoperiodic changes in day-length and is associated with mood disorders (17). The Global Seasonality Score (GSS) was calculated using the Seasonal Pattern Assessment Questionnaire (18). The GSS is composed of six items assessing seasonal change across sleep length, social activity, mood, weight, appetite and energy. Each item is scored from *0 (“no change”)* to *4 (“extremely marked change”)*. Items are summed to calculate the GSS. Following validated cut-offs (19), a GSS ≥11 was coded as having seasonality.
- <u>Social jetlag:</u> Social jetlag represents the mismatch between biological time and social time and is a marker of circadian disturbance (20). First, sleep midpoint, the halfway point (clock time) between sleep onset and waking up, was calculated on free (non-working) days and on working days. Next, the difference between sleep midpoint on free days and on working days was calculated. Given the wide age range in AGDS (18-90 years) and the fact that sleep timing changes with age, age-adjusted thresholds for social jetlag were applied, corresponding to the average sleep midpoint of the age group +1 standard deviation (SD):

- 18-24 year-olds: sleep midpoint on free days ≥2.5-hours later than on working days.
- 25-34 year-olds: sleep midpoint on free days ≥2.25-hours later than on working days.
- 35-64 year-olds: sleep midpoint on free days ≥2-hours later than on working days.
- 65+ year-olds: sleep midpoint on free days ≥1.25-hour later than on working days.
- <u>Evening chronotype</u>: Chronotype is the genetically-influenced preference for the daily timing of sleep, wake, and other phenomena, and is associated with mood disorders (21, 22). Chronotype was defined using item 19 of the Morningness-Eveningness Questionnaire (MEQ) (23). Those reporting being “*Definitely an “evening” type”* were coded as having an evening chronotype.
- <u>Delayed sleep midpoint:</u> Delayed sleep midpoint is an expression of evening chronotype (which is the *preference* for sleep-wake timing) and is genetically influenced (24) but also influenced by behaviour (e.g., work schedules). First, sleep midpoint on free (non-working) days was calculated. Then, age-adjusted cut-offs, corresponding to average sleep midpoint of the age group + 1 SD, were applied to determine delayed sleep midpoint:

- 18-24-year-olds: sleep midpoint later than 6am;
- 25-34-year-olds: sleep midpoint later than 5:30am;
- 35-64-year-olds: sleep midpoint later than 5am; and
- 65+ year-olds: sleep midpoint later than 4:30am.
- <u>Hypersomnia:</u> Hypersomnia is prevalent symptom of mood disorders and may plausibly stem from circadian disruption (e.g., weakened circadian signal of alertness leading to increased sleepiness). Using the DSM-5 MDD criteria, those reporting to *“sleep much more than usual”* during the period in which their feelings of depression were the worst were coded as having hypersomnia.
- <u>Sleep inertia:</u> Sleep inertia is proposed to be a marker of waking during the biological (circadian) night. Sleep inertia was defined using item 8 from the MEQ (23). Those reporting to be “*very tired”* in the first half-hour after waking in the morning were coded as having sleep inertia.

Table 2 reports the numbers and proportions of participants meeting these circadian criteria. The sum of all criteria was computed and those meeting ≥3 of 6 criteria were coded as the *“circadian subtype”* group; those meeting fewer than 3 were coded as the *“non-circadian group”*.

**Table 1.** Demographic and clinical characteristics of the analytic sample.

| Mean (SD) | Total sample<br>N=11,380 | Circadian subtype<br>N=2,604 (23%) | Non-circadian subtype<br>N=8,776 (77%) | *p-value* |
| --- | --- | --- | --- | --- |
| **Age** | 43.80 (15.28) | 37.87 (13.62) | 45.56 (15.30) | <0.001 |
| **Sex; N (%)** |  |  |  | <0.001 |
| Female | 8,509 (75) | 2,090 (80) | 6,419 (73) |  |
| Male | 2,861 (25) | 513 (20) | 2,348 (27) |  |
| Information not provided | 10 (<1) | 1 (<1) | 9 (<1) |  |
| **BMI** | 28.49 (7.28) | 29.66 (8.06) | 28.15 (6.99) | <0.001 |
| **Marital status, N (%)** |  |  |  | <0.001 |
| Married or de facto | 6,183 (54) | 1,102 (42) | 5,081 (58) |  |
| Separated or divorced | 1,676 (15) | 373 (14) | 1,303 (15) |  |
| Widowed | 192 (2) | 25 (1) | 167 (2) |  |
| Never married | 3,311 (29) | 1,102 (42) | 2,209 (25) |  |
| Information not provided | 18 (<1) | 2 (<1) | 16 (<1) |  |
| **Highest level of education, N (%)** |  |  |  | <0.001 |
| Degree | 4,047 (36) | 1,014 (40) | 3,033 (35) |  |
| Postgraduate | 3,194 (28) | 602 (23) | 2,592 (29) |  |
| Certificate or diploma | 2,637 (23) | 638 (24) | 1,999 (23) |  |
| Senior high school (Y 11-12) | 870 (8) | 226 (9) | 644 (7) |  |
| Junior high school or less (Y 1-10) | 623 (5) | 122 (4) | 501 (6) |  |
| No formal education | 4 (<1) | 1 (<1) | 3 (<1) |  |
| Information not provided | 5 (<1) | 1 (<1) | 4 (<1) |  |
| **Meeting MDD Criteria, N “Yes” (%)** | 10,840 (95) | 2,523 (97) | 8,317 (95) | <0.001 |
| **Weight gain during episode, N “Yes” (%)** | 3,815 (36) | 1,061 (41) | 2,754 (31) | <0.001 |
| **Depressive episodes, N (%)** |  |  |  | <0.001 |
| 1-2 episodes | 1,275 (11) | 182 (7) | 1,093 (12) |  |
| 3-4 episodes | 2,301 (20) | 445 (17) | 1,856 (21) |  |
| 5-6 episodes | 1,655 (15) | 400 (15) | 1,255 (14) |  |
| 7-9 episodes | 579 (5) | 159 (6) | 420 (5) |  |
| 10 or more episodes | 4,529 (40) | 1,354 (52) | 3,175 (36) |  |
| Information not provided | 1,041 (9) | 64 (2) | 977 (11) |  |
| **Age of onset** | 22.09 (11.26) | 18.68 (8.40) | 23.20 (11.84) | <0.001 |
| **Mania (ASRM)** | 2.58 (2.09) | 2.99 (2.06) | 2.46 (2.08) | <0.001 |
| **Psychosis (CAPE)** | 0.87 (1.34) | 1.18 (1.54) | 0.78 (1.26) | <0.001 |
| **Suicidality (SIDAS)** | 5.69 (9.27) | 8.18 (10.65) | 4.96 (8.68) | <0.001 |
| **Psychological distress (SPHERE)** | 3.40 (3.69) | 5.40 (3.72) | 3.56 (3.57) | <0.001 |
| **Somatic complaints (SPHERE)** | 4.61 (3.49) | 6.33 (3.43) | 4.10 (3.34) | <0.001 |
| **Atypical depression, N “Yes” (%)** | 2,344 (21) | 940 (36) | 1,404 (16) | <0.001 |
| **Bipolar disorder, N “Yes” (%)** | 1,006 (9) | 309 (12) | 697 (8) | <0.001 |
| **Medication use, N taking (%)** |  |  |  |  |
| SSRI | 9,333 (82) | 2,251 (86) | 7,082 (81) | <0.001 |
| SNRI | 5,536 (49) | 1,421(55) | 4,115 (47) | <0.001 |
| TCA | 2,778 (24) | 673 (26) | 2,105 (24) | 0.056 |
| Lithium | 764 (7) | 193 (7) | 571 (7) | 0.115 |

*Notes.* ASRM: Altman Self-Rating Mania Scale; CAPE: Community Assessment of Psychic Experiences; SIDAS: Suicidal Ideation Attributes Scale; SPHERE: Somatic and Psychological Health Report; SNRI: Serotonin-norepinephrine reuptake inhibitor; SSRI: Selective serotonin reuptake inhibitors; TCA: Tricyclic antidepressants. Comparisons of clinical scores accounted for the effect of age and sex.

**Table 2.** Circadian features in the whole sample (N=11,380)

|  | Variable used | Age group | Mean (SD) | Cut-off / category used | N (%) meeting criteria |
| --- | --- | --- | --- | --- | --- |
| **Seasonality** | SPAQ GSS |  | 6.59 (4.69) | GSS > 11 | 2,190 (19.2) |
| **Social jetlag** | Weekend-Weekday<br>difference in<br>sleep midpoint | 18-24 | 1.33 (1.11) | > 2h30min | 1,390 (12.2) |
|  |  | 25-34 | 1.23 (1.01) | > 2h15min |  |
|  |  | 35-64 | 0.95 (0.86) | > 2h00min |  |
|  |  | 65+ | 0.46 (0.69) | > 1h30min |  |
| **Evening chronotype** | MEQ item 19 |  |  | “Definitely an “evening” type” | 2,826 (24.8) |
| **Delayed sleep midpoint** | Sleep midpoint | 18-24 | 28.49 (1.36) | > 6:00am | 1,409 (12.4) |
|  |  | 25-34 | 28.00 (1.35) | > 5:30am |  |
|  |  | 35-64 | 27.48 (1.29) | > 5:00am |  |
|  |  | 65+ | 27.17 (1.18) | > 4:30am |  |
| **Hypersomnia** | DSM-5 criteria |  |  | Reported sleeping more<br>during worst depressive episode | 5,831 (51.2) |
| **Sleep inertia** | MEQ item 8 |  |  | “Very tired” | 4,391 (38.6) |

Notes. SPAQ: Seasonal Pattern Assessment Questionnaire; GSS: General Seasonality Score; MEQ: Morningness-Eveningness Questionnaire.

### c. Assessments

#### Clinical variables

Information about demographics, age of onset, and clinical course were assessed by self-report items.

- Hypo/manic-like experiences were assessed using items adapted from the Altman Self-Rating Mania Scale (ASRM) (25). The ASRM is 5-item self-report tool designed to assess the presence and severity of manic symptoms in domains including mood, self-confidence, sleep disturbances, speech, and activity level over a week. The total score was used (range 0-20).
- Psychotic-like experiences were assessed using items adapted from the Community Assessment of Psychic Experiences (CAPE) (26). The CAPE is a 15-item self-report questionnaire using to assess psychotic-like experience in the community, focusing on persecutory ideation, bizarre experiences, and perceptual abnormalities. The total score was used (range 15-75).
- Psychological and somatic distress were assessed using the Somatic and Psychological Health Report (SPHERE) (27). The SPHERE is a 12-item self-rated tool developed to screen for anxiety, depression, and somatisation in primary care. Sub-scale scores for psychological health (PSYCH) and for physical symptoms and fatigue (SOMA) were derived and used separately (ranges 0-6).
- Suicidal ideation was assessed with the Suicidal Ideation Attributes Scale (SIDAS) (28). The SIDAS is a 5-item scale measuring frequency and severity of suicidal ideation in the past month. Each item is scored on a scale from “*0 (Never)*” to “*10 (Always)*”. The total score was used (range 0-50).

#### Antidepressants, efficacy and sides effects

Participants’ subjective experiences with 10 of the most commonly prescribed antidepressants in Australia at the time of the survey were assessed. Efficacy was rated on an ordinal scale (0 = *not at all well*; 1 = *moderately well*; 2 = *very well*), excluding “*I don’t know*” responses, and compared across three major classes: SSRIs (sertraline, escitalopram, citalopram, fluoxetine, and paroxetine), SNRIs (venlafaxine, desvenlafaxine, and duloxetine), and tricyclic antidepressants (TCAs; amitriptyline and mirtazapine). In the case of different ratings for antidepressants within the same class, the highest efficacy rating was used as the class-level rating. Side effects while taking antidepressants were assessed as a binary variable (0=no, 1=yes) across 25 possible symptoms, including rash, runny nose, muscle pain, constipation, diarrhea, drowsiness, agitation, fatigue, suicidal thoughts, dry mouth, blurred vision, difficulty getting to sleep, weight gain, sweating, vomiting, shaking, increased anxiety, attempted suicide, headache, reduced sexual desire, dizziness, nausea, weight loss, and other side effects. Participants reported side effects only for antidepressants they had taken.

### d. Polygenic risk scores

DNA samples were collected using saliva kits, genotyped using the Illumina Global Screening Array V.2.0, and processed using standardised quality control (QC) measures. Pre-imputation QC was done using PLINK 1.9 (29, 30) and involved removing single nucleotide polymorphisms (SNPs) with a minor allele frequency <0.005, a SNP call rate <97.5%, and Hardy-Weinberg equilibrium (p<1x10-6). Genotypes were imputed using 1000 Genomes, HRC v1.1, and TopMed r2 reference panel (31). Ancestry was inferred using the first three principal components (PCs), by projecting PCs on 1000 Genomes data using GCTA (32). The samples within six standard deviations (SD) of the PCs of each ancestry in 1000 Genomes are assigned as the same ancestry. The latest publicly available genome-wide association study (GWAS) results for mental disorders and sleep, circadian, metabolic, and inflammatory traits were used as weights in the calculation of each PGS (see Table S1). Where applicable, leave-one-out summary statistics were used for GWAS studies that included AGDS participants to avoid over-estimation. SBayesRC was used to generate allele weights for each PGS (33). The posterior SNP effects for each disorder/trait were used to generate PGS for each participant using the PLINK score function (29). Each PGS was standardised with the sample. Effect sizes are interpretable per SD unit increase in the AGDS PGS.

### e. Statistical analyses

Analyses were conducted in RStudio (R version 4.3.0) (34). Demographic and clinical characteristics of circadian vs non-circadian depressive profiles were compared using Welch’s t-test for age, Analysis of Covariance (adjusting for age/sex) for continuous variables and Pearson’s Chi-squared (χ²) test for categorical variables.

Logistic regression was used to estimate (1) associations between 27 PGS (mental health, physical health, and circadian and sleep-related PGS) and the circadian subtype, adjusting for age, sex, and the first four genetically-inferred ancestry PCs; and (2) associations between antidepressants efficacy and side effects and circadian subtype, adjusting for age and sex. To control for multiple testing, Bonferroni correction was applied at the following threshold and significance level, *p*=[0.05/27]=0.0019, accounting for six mental health PGS, 16 physical health PGS, and five sleep and circadian-related PGS. For antidepressants efficacy, the Bonferroni threshold was 0.017 (*p*<0.05/3) and for side effects, the Bonferroni threshold was 0.002 (*p*<0.05/25). Associations between each binary circadian criterion and the 27 selected PGS and antidepressant efficacy are reported in Figures S6-S9.

## 3. Results

### a. Distribution of circadian criteria

Of 11,380 participants with non-missing data for ≥3 circadian criteria, a subset of 2,604 participants (80% female; Mean age = 37.87±13.62) were classed as having a *circadian subtype.* Hypersomnia was the most frequently endorsed criterion in participants with the circadian subtype (N=2,182; 84%), followed by sleep inertia (N=2,143; 83%), evening chronotype (N=1,774; 68%), delayed sleep midpoint (N=1,099; 42%), and seasonality (N=1,071; 41%). The least commonly endorsed criterion was social jetlag (N=914; 35%). The most common combinations of these criteria were hypersomnia, sleep inertia, and evening chronotype (N=481; 18%). The distributions of these criteria are shown in Figure 1.

**Figure 1.**
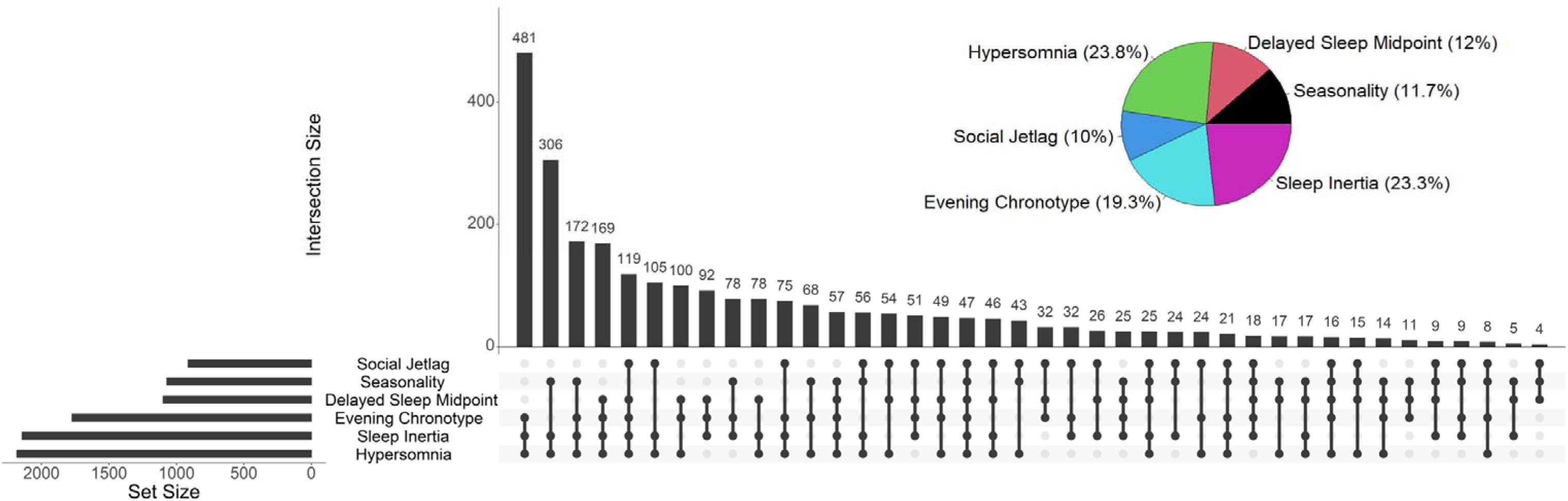
Distribution of circadian criteria in participants with the circadian subtype of depression (N=2,604). UpSet plot of intersection between circadian criteria. Each row represents on circadian criterion, and each column indicates a specific combinations of criteria. Filled dots denote which criteria are included in each intersection. Bar height above each column shows the number of participants meeting that combination of criteria. Pie chart illustrates the relative proportion of each circadian criteria.

### b. Demographics and clinical characteristics of the circadian subtype

As shown in Table 1, compared to the non-circadian subtype of depression (N=8,776), circadian cases were younger (37.87±13.62 vs. 45.56±15.30; *p*<0.001) and more often female (80% vs 73%, *p*<0.001). They reported an earlier age of onset of depression (18.68±8.40 vs. 23.20±11.84, *p*<0.001), greater severity of hypo/manic-like experiences (2.99±2.06 vs. 2.46±2.08, *p*<0.001), psychotic-like experiences (1.18±1.54 vs. 0.78±1.26, *p*<0.001), suicidality (SIDAS) (8.18±10.65 vs. 4.96±8.68, *p*<0.001), psychological distress (5.40±3.72 vs. 3.56±3.57, *p*<0.001), and somatic complaints (6.33±3.43 vs. 4.10±3.34, *p*<0.001).

### c. Genetic profile of the circadian subtype

#### i. PGS for mental disorders

As shown in Figure 2A and Table S2, the circadian subtype was associated with higher genetic risk (PGS) for attention-deficit/hyperactivity disorder (ADHD) (odds ratio [OR]=1.11; 95%CI=[1.06, 1.17]; *p*<0.001), major depression (OR=1.11 [1.06, 1.16]; *p*<0.001), and BD (OR=1. 09 [1.04, 1.14]; *p*<0.001). The circadian subtype was also associated with higher schizophrenia PGS (OR=1.07 [1.02, 1.13]; *p*=0.006), however this association did not survive Bonferroni correction. Neither the neuroticism (OR=0.98 [0.93, 1.03]; *p*=0.419) or autism PGS (OR=1.04 [0.99, 1.09]; *p*=0.105) were associated with the circadian subtype.

**Figure 2.**
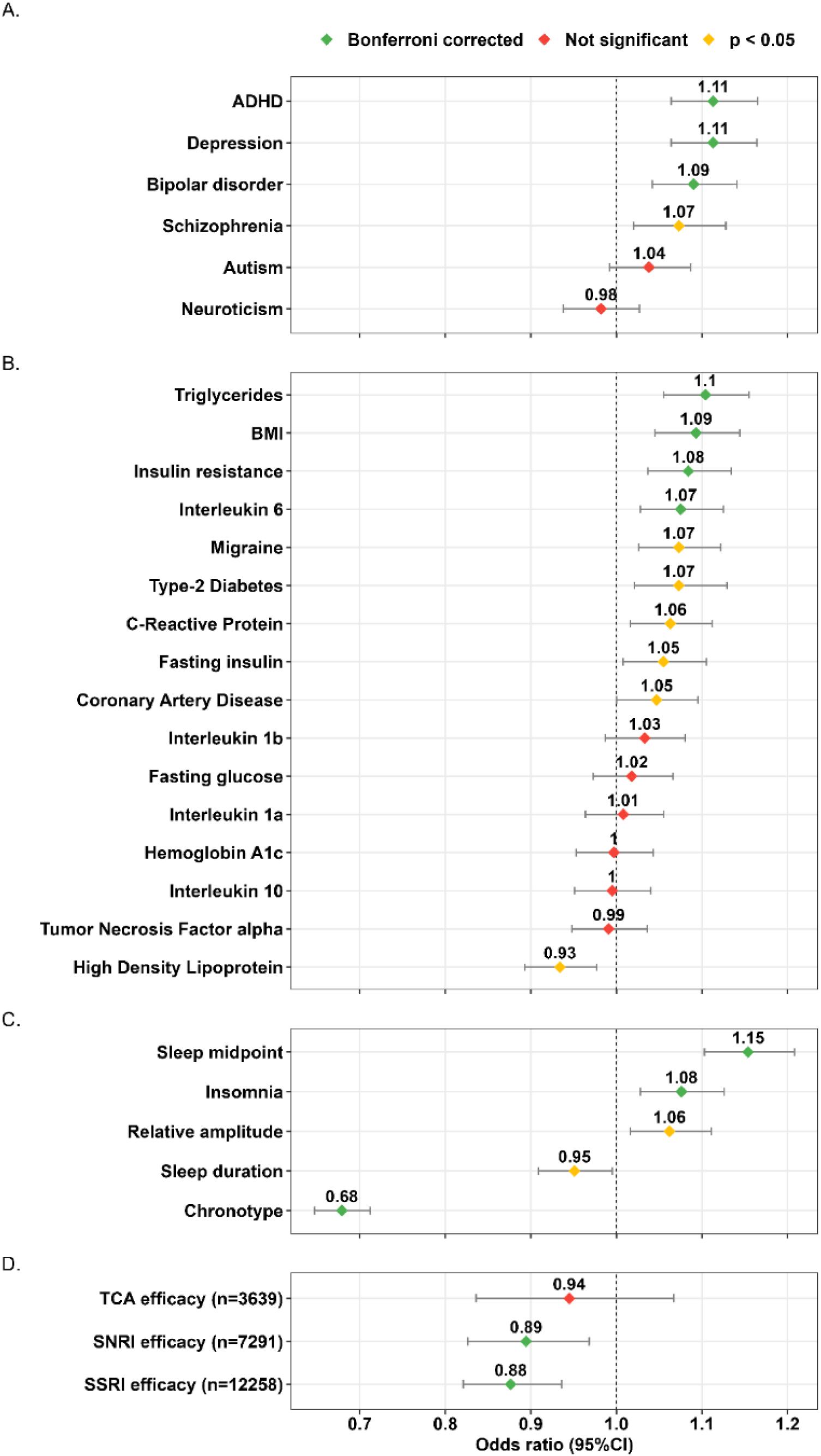
Associations between the circadian subtype and (A) polygenic risk scores (PGS) for mental disorders; (B) PGS for physical health measures (metabolic and inflammatory-related markers); (C) PGS for sleep and circadian-related measures, and (D) treatment response (n=2,604 [23%] with circadian cases; n=8,776 [77%] with non-circadian cases). *Notes.* Results shown are separate logistic regression models for each variable with the circadian subtype, with covariates of age and sex for all models, and additionally the first 4 genetically inferred ancestry PCs for PGS models. Significance levels: *Green*, Bonferroni-corrected (at alpha level p<0.0019 [0.05/27] 6 mental disorder PGS, 16 physical health PGS, 5 sleep and circadian-related measures PGS), and at alpha level p<0.0167 [0.05/3] for treatment response); *Yellow*, Uncorrected (*p*<0.05); *Red*, Not significant. Error bars represent 95% confidence interval. ADHD: Attention-Deficit Hyperactivity Disorder; BMI: Body Mass index; SNRI: Serotonin-norepinephrine reuptake inhibitor; SSRI: Selective serotonin reuptake inhibitors; TCA: Tricyclic antidepressants.

#### ii. PGS for physical health

As shown in Figure 2B and Table S2, the circadian subtype was associated with higher PGS for BMI (OR=1.09 [1.05, 1.14]; *p*<0.001), triglycerides (OR=1.10 [1.06, 1.16]; *p*<0.001), IL-6 (OR=1.08 [1.03, 1.13]; *p*=0.001), and insulin resistance (OR=1.08 [1.04, 1.13]; *p*<0.001). The circadian subtype was also associated with lower PGS for high-density lipoprotein (HDL) (OR=0.93 [0.89, 0.98]; *p*=0.003) and higher PGS for migraine (OR=1.07 [1.03, 1.12]; *p*=0.002), C-reactive protein (CRP) (OR=1.06 [1.02, 1.11]; *p*=0.008), type 2 diabetes (OR=1.07 [1.02, 1.13]; *p*=0.006), coronary artery disease (OR=1.05 [1.00, 1.10]; *p*=0.048) and fasting insulin (OR=1.06 [1.01, 1.11]; *p*=0.023), however these associations did not survive Bonferroni correction. The PGS for interleukin-10 (IL-10), IL-1β, IL-12, fasting glucose, tumour necrosis factor alpha (TNF-α), and glycated haemoglobin (HbA1c) were not associated with the circadian subtype (all *p*>0.159).

#### iii. PGS for sleep and circadian traits

As shown in Figure 2C and Table S2, the circadian subtype was associated with lower PGS for chronotype (i.e., higher liability for eveningness) (OR=0.68 [0.65, 0.71]; *p*<0.001) and higher PGS for insomnia (OR=1.08 [1.03, 1.13]; *p*=0.002) and sleep midpoint (i.e., higher liability for later sleep midpoint) (OR=1.16 [1.10, 1.21]; *p*<0.001). The circadian subtype was also associated with higher PGS for low relative amplitude (i.e., higher liability for weaker rest-activity amplitude) (OR=1.06 [1.02, 1.11]; *p*=0.008) and lower sleep duration (OR=0.95 [0.91, 0.99]; *p*=0.029), however these associations did not survive Bonferroni correction.

### d. Treatment response outcomes

Finally, as shown in Figure 2D and Table S2, the circadian subtype was associated with lower efficacy of SSRIs (OR=0.88 [0.82, 0.94]; *p*<0.001 and SNRIs (OR=0.89 [0.83, 0.97]; *p*=0.006), but not with efficacy of TCAs (OR=0.94 [0.84, 1.07]; *p*=0.362). Within-class comparisons of antidepressant efficacy are reported in the Supplementary Materials. The circadian subtype was associated with a broad profile of self-reported antidepressant side effects, including runny nose, muscle pain, blurred vision, constipation, drowsiness, diarrhea, fatigue, difficulty sleeping, sweating, agitation, attempted suicide, shaking, dry mouth, headache, weight gain, suicidal thoughts, anxiety, reduced sexual desire, dizziness, and nausea. No significant association was found with reporting “rash”, “other side effects”, “no side effects”, and “weight loss” (Figure S2 and Table S4).

## 4. Discussion

In a large genetically-informative cohort of adults with severe, recurrent forms of depressive disorders, 23% of participants were classified as having a ‘circadian subtype’ of depression. These participants were younger, more commonly female, had an earlier age of onset and reported a more severe clinical profile in terms of the number of depressive episodes and severity of multiple symptom dimensions. The circadian subtype was associated with lower reported efficacy of SSRIs and SNRIs and more side effects, suggestive of its clinical utility. As greater depression severity is typically considered to be a predictor of enhanced response to antidepressant medications (2, 3), our finding of the opposite may be of clinical significance and supportive of the hypothesis of this circadian subtype being underpinned by a unique pathophysiological mechanism from other forms of severe or persistent depressive disorders. Genetically, circadian cases had higher genetic risk for some neurodevelopmental conditions (ADHD) and mood disorders (major depression and BD); sleep-wake cycle phenotypes including delayed sleep midpoint, insomnia, and evening preference; and metabolic (insulin resistance, BMI, triglycerides) and inflammatory (IL-6) markers. These clinical, treatment response and genetic findings support the notion that a circadian subtype represents a distinct and clinically-relevant profile of depressive disorders.

The increased genetic risk for ADHD, major depression and BD in circadian cases warrants further exploration. Delayed phase of circadian markers, such as dim light melatonin onset, sleep-wake timing, and evening chronotype are common in ADHD (35). Specific circadian SNPs have been linked to ADHD symptoms (36) and genetic risk for dysregulated melatonin appears to be correlated with ADHD (37). Additionally, another study conducted in AGDS observed an association between the ADHD PGS and increased likelihood of having an earlier age of onset, greater recurrence of depressive episodes, and atypical depression (38). Circadian disruption is also a hallmark of BD (39), often preceding the onset of episodes (17), and is potentially implicated in the switch between depressive and manic states in BD (40). Shared genetic pathways further support circadian involvement in BD, with dysregulated clock genes in BD (41) and BD polygenic risk predicting later timing of circadian activity peaks (13). In addition to higher PGS for BD, we also observed a higher enrichment for self-reported lifetime diagnoses of BD in the circadian cases but note that almost 700 cases of BD were assigned to the non-circadian cases.

The circadian subtype was also associated with genetic risk for metabolic and inflammatory markers. This aligns with evidence linking circadian disruption to metabolic dysregulation and chronic inflammation (42), notably, adolescents with delayed sleep wake cycle showing metabolic abnormalities (43) and insulin resistance being linked to abnormal melatonin-sleep timing (44, 45). Circadian disruption may contribute to cardiometabolic dysfunction (46), with genetic vulnerability in metabolic and inflammatory pathways worsening outcomes and potentially leading to treatment resistance. The circadian subtype was also associated with higher PGS for later sleep midpoint, insomnia, and evening chronotype, which provides suggestive evidence that the circadian subtype captures a biologically meaningful subset, as opposed to these features being epiphenomena. Clinically, circadian cases reported lower SSRI and SNRI efficacy and more side effects, consistent with evidence that circadian misalignment may respond less favourably to standard pharmacotherapies (15, 47) or the notion that conventional antidepressants do not adequately correct sleep-wake/circadian disturbance.

Other proposed subtypes of depression, such as atypical depression, immuno-metabolic depression, and bipolar depression share some related features with the circadian subtype of depression. Notably, atypical depression, defined by mood reactivity, weight gain, somatic symptoms, and hypersomnia and being initially linked to preferential response to monoamine oxidase inhibitors (48), also shows poorer response to SSRIs and SNRIs and includes sleep-related features (i.e. hypersomnia) in its definition. However, its strong reliance on weight gain during episodes or over time as a criterion is likely to conflate heterogeneous and potentially unrelated factors, including anxiety and neuroticism, alongside circadian characteristics. We note that while our circadian subtype is enriched for cases who also meeting criteria for atypical depression, ∼1400 cases with atypical depression were assigned to the non-circadian subtype group (Table 1). This indicates that these phenotypes are partly overlapping but separable constructs. In another AGDS study (49) we showed that restricting the definition of atypical depression to reliably reported features (i.e., weight gain and hypersomnia) demonstrated important differences with the circadian subtype, notably higher PGS for neuroticism (full comparison reported in Table S5). By contrast, our circadian phenotype is defined specifically by 24-hour sleep-wake and circadian-related markers, arguably providing stronger specificity to circadian disturbance as an underlying pathophysiology. Similarly, immuno-metabolic depression is characterised by a cluster of energy-related depressive symptoms and related biomarkers, including systemic low-grade inflammation and metabolic abnormalities (50). Immune-metabolic depression is also characterised by a poor response to standard pharmacology and evidence suggests intervention targeting inflammation, metabolism, or lifestyle factors may prove more effective (51). However, immune-metabolic depression may result from a range of different and confounding factors, including age, chronicity of illness, weight gain, medication exposure, and other comorbidities (52). Thus, the clinical utility of immune-metabolic depression in the early identification and intervention of emerging depressive disorders is limited.

Circadian disruptions are also reported in bipolar depression (53), alongside mixed efficacy of SSRIs and SNRIs (54) and probable metabolic-inflammatory factors (55). However, around half of BD cases initially present with depressive episodes that are difficult to distinguish from unipolar depression, with (hypo-)manic episodes often emerging after several depressive episodes (56). This renders the early identification of bipolar depression particularly challenging (57). Other common subtypes of depression include treatment-resistant depression, with limited utility to guide preferential first treatment priority by definition, and melancholic depression, which can be regarded as a marker of severe or difficult-to treat MDD conditions (58). Overall, distinctions between depressive phenotypes are key to inform different treatments approaches, notably first-line interventions. Compared with other depressive subtypes, our proposed circadian subtype of depression demonstrates clear genetic, clinical, and treatment (self-rated) correlates, which support the formal evaluation of a stratification approach based on circadian factors to guide early intervention strategies. Additionally, circadian disturbances have been associated with transition to more severe clinical staging, suggesting its predictive value for illness progression in early intervention (59).

## Limitations

Several limitations need to be acknowledged. First, the data were collected through a self-report survey, which while allowing richer access to the lived experience of depression, may be subject to biases (e.g., recall biases). Second, the data presented were cross-sectional, rendering causal inferences challenging. Third, while the 10 most common antidepressants were recorded, their classification in broader categories may mask important sources of heterogeneity. Fourth, individuals in this sample are limited to genetically-inferred European ancestry, which may limit the generalisability of these findings. Finally, while the self-reported nature of these circadian criteria is an advantage for investigation in large-scale studies, these criteria do not directly capture internal physiological circadian markers (e.g., phase, amplitude, misalignment).

## Conclusion

This study strengthens the evidence of a circadian subtype of depression as a valid and clinically useful phenotype of depression associated with differential treatment response, clinical course characteristics, and genetic profiles. Future studies are needed to replicate our findings in diverse samples, to investigate whether the phenotype has predictive utility, and to explore whether the circadian subtype indeed preferentially captures cases with circadian disruption as a major pathophysiology.

## Data Availability

All data produced in the present study are available upon reasonable request to the authors

## Acknowledgements

We are indebted to all of the Australian Genetics of Depression Study participants for giving their time to contribute to this study. We thank all the people who helped in the conception, implementation, beta testing, media campaign, and data cleaning. We would like to thank the research participants and employees of 23andMe Research Institute for making this work possible. The Australian Genetics of Depression Study was primarily funded by grant 1086683 from the National Medical Health and Research Council (NHMRC) of Australia. This work was further supported by philanthropic donations from families who are affected by mental illness (who would like to be left anonymous) awarded to MS; NHMRC EL1 Investigator Grants awarded to JJC and BLM (GNT2008196 and GNT2017176, respectively); a Wellcome Trust Mental Health Award (227089/Z/23/Z); NHMRC L1 and L2 Investigator Grants (APP117291, APP2025674) awarded to SEM; and an NHMRC L3 Investigator Grant (GNT2016346) awarded to IBH.

