## Supplementary Materials for "Clinical and genetic correlates of a circadian subtype of depression in the Australian Genetics of Depression Study"

**Figure S1	Flow chart of participants selection included in this study**

**Table S1	Demographic and clinical characteristics of included versus not included participants.**

**Table S2	Genome-wide association studies used for PGS calculation**

**Figure S2	Number of circadian criteria met in the whole sample (N=11,380)**

**Table S3	Associations between the circadian subtype and polygenic risk scores and reported antidepressants side effects**

**Table S4	Sensitivity analyses, correcting for weight gain effect**

**Figure S3	Associations between the circadian subtype and self-reported side effects of antidepressants (n=2,604 with circadian cases; n=8,776 with non-circadian cases)**

**Table S5	Associations between the circadian subtype and reported antidepressant side effects**

**Table S6	Comparison between atypical depression and circadian subtype of depression in AGDS**

**Figure S4	Associations between number of criteria and clinical variables.**

**Figure S5	Associations between the circadian subtype and polygenic risk scores for mental disorders, physical health, and sleep and circadian factors and treatment response restricted to those with data complete for all 6 circadian criteria.**

**Table S7	Associations between the circadian subtype and polygenic risk scores and reported antidepressants side effects restricted to those with data complete for all 6 circadian criteria (n=2,180 with circadian cases; n=6,548 with non-circadian cases)**

**Figure S6	Associations between each binary circadian criteria and polygenic risk scores for mental disorders.**

**Figure S7	Associations between each binary circadian criteria and polygenic risk scores for physical health.**

**Figure S8	Associations between each binary circadian criteria and polygenic risk scores for sleep and circadian factors.**

**Figure S9	Associations between each binary circadian criteria and self-reported antidepressant efficacy.**

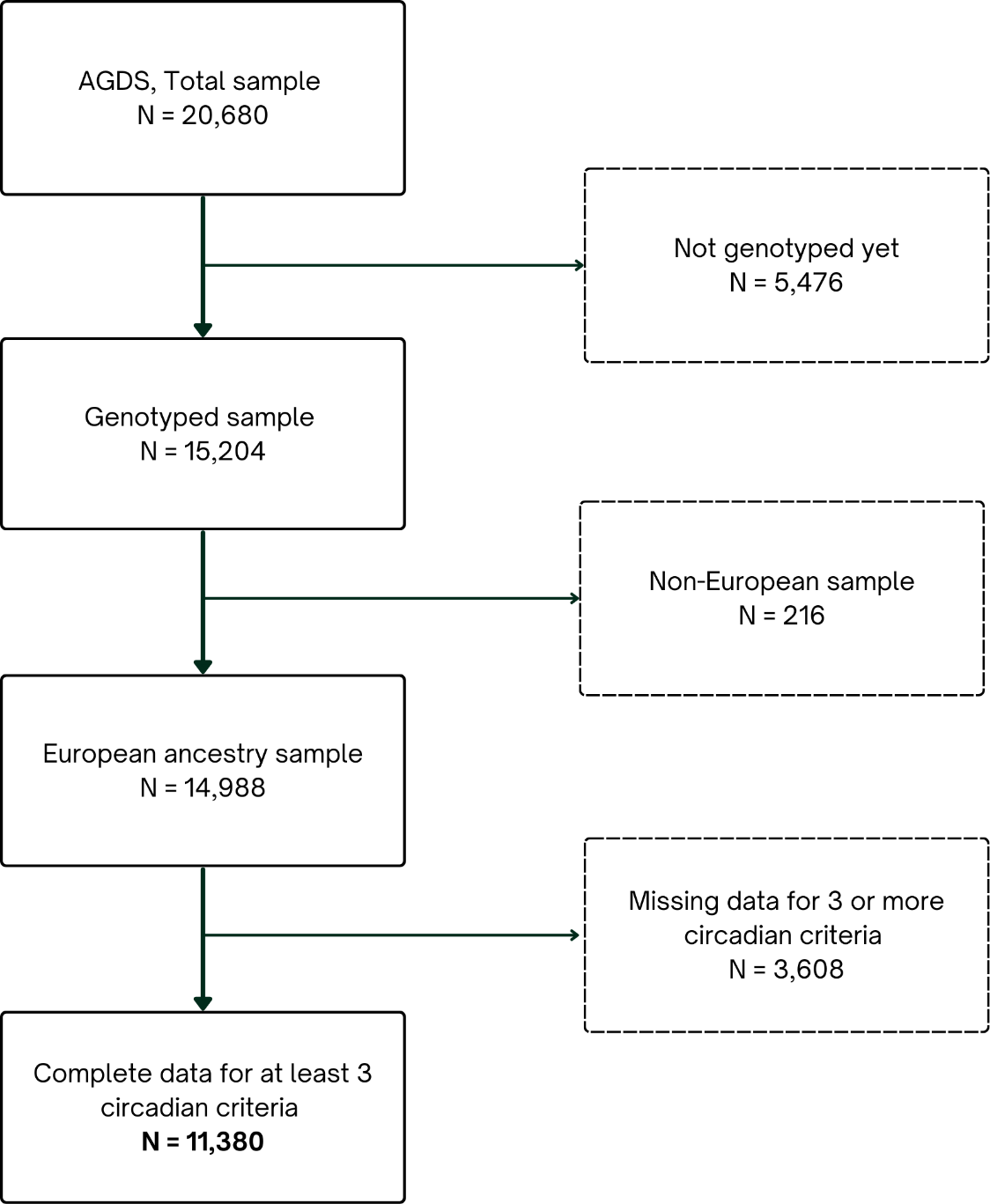
**Figure S1 Flowchart of participants selection for this study.** European ancestry was determined using a cut-off of 6 standard deviation above or below the sample mean of PC1 or PC2.

| **Table S1. Demographic and clinical characteristics of included versus not included participants.** | | | |
| --- | --- | --- | --- |
| Mean (SD) | **AGDS sample included; N=11,380 (68%)** | **AGDS sample not included; N=3,608 (32%)** | ***p-value*** |
| **Age** | 43.80 (15.28) | 43.54 (15.27) | 0.365 |
| **Sex; N (%)** |  |  | 0.881 |
| Female | 8,509 (75) | 2,692 (75) |  |
| Male | 2,861 (25) | 912 (25) |  |
| Information not provided | 10 (<1) | 4 (<1) |  |
| **BMI** | 28.49 (7.28) | 28.42 (7.35) | 0.621 |
| **Marital status**, N (%) |  |  | 0.015 |
| Married or de facto | 6,183 (54) | 1,903 (53) |  |
| Separated or divorced | 1,676 (15) | 587 (16) |  |
| Widowed | 192 (2) | 65 (2) |  |
| Never married | 3,311 (29) | 1,039 (29) |  |
| Information not provided | 18 (<1) | 14 (<1) |  |
| **Highest level of education**, N (%) |  |  |  |
| Degree | 4,047 (36) | 1,161 (32) | <0.001 |
| Postgraduate | 3,194 (28) | 950 (26) |  |
| Certificate or diploma | 2,637 (23) | 911 (25) |  |
| Senior high school (Y 11-12) | 870 (8) | 310 (9) |  |
| Junior high school or less (Y 1-10) | 623 (5) | 240 (7) |  |
| No formal education | 4 (<1) | 2 (<1) |  |
| Information not provided | 5 (<1) | 34 (10) |  |
| **Meeting MDD Criteria**, N “Yes” (%) | 10,840 (95) | 3,415 (95) | 0.155 |
| **Weight gain during episode**, N “Yes” (%) | 3,815 (36) | 1,222 (34) | 0.454 |
| **Depressive episodes**, N (%) |  |  | 0.309 |
| 1-2 episodes | 1,275 (11) | 355 (10) |  |
| 3-4 episodes | 2,301 (20) | 729 (20) |  |
| 5-6 episodes | 1,655 (15) | 516 (14) |  |
| 7-9 episodes | 579 (5) | 194 (5) |  |
| 10 or more episodes | 4,529 (40) | 1,438 (40) |  |
| Information not provided | 1,041 (9) | 376 (10) |  |
| **Age of onset** | 22.09 (11.26) | 22.28 (11.35) | 0.328 |
| **Mania (ASRM)** | 2.58 (2.09) | 2.80 (2.08) | <0.001 |
| **Psychosis (CAPE)** | 0.87 (1.34) | 0.97 (1.40) | <0.001 |
| **Suicidality (SIDAS)** | 5.69 (9.27) | 5.80 (9.19) | 0.541 |
| **Psychological distress (SPHERE)** | 3.40 (3.69) | 4.04 (3.65) | 0.592 |
| **Somatic complaints (SPHERE)** | 4.61 (3.49) | 4.70 (3.55) | 0.368 |
| **Atypical depression**, N “Yes” (%) | 2,344 (21) | 774 (21) | 0.155 |
| **Bipolar disorder**, N “Yes” (%) | 1,006 (9) | 331 (9) | 0.562 |
| **Medication use**, N taking (%) |  |  |  |
| SSRI | 9,333 (82) | 2,925 (81) | 0.210 |
| SNRI | 5,536 (49) | 1,755 (49) | 1 |
| TCA | 2,778 (24) | 861 (24) | 0.518 |
| Lithium | 764 (7) | 237 (7) | 0.781 |
| *Notes.* ASRM: Altman Self-Rating Mania Scale; CAPE: Community Assessment of Psychic Experiences; SIDAS: Suicidal Ideation Attributes Scale; SPHERE: Somatic and Psychological Health Report; SNRI: Serotonin-norepinephrine reuptake inhibitor; SSRI: Selective serotonin reuptake inhibitors; TCA: Tricyclic antidepressants. Comparisons of clinical scores accounted for the effect of age and sex. | | | |

| **Table S2 Genome-wide association studies used for PGS calculation** | | | | |
| --- | --- | --- | --- | --- |
| **PGS** | **GWAS used to derived PGS** | **LOO** | **N cases** | **N controls** |
| **Mental disorders/conditions** | | | | |
| ADHD | (Demontis et al., 2023) |  | 38,691 | 186,843 |
| Depression | (Adams, 2025) | Y | 688,808 | 4,364,225 |
| Bipolar disorder | (O’Connell et al., 2025) | Y | 158,036 | 2,796,499 |
| Schizophrenia | (Trubetskoy et al., 2022) | N | 76,755 | 243,649 |
| Autism | (Grove et al., 2019) |  | 18,381 | 27,969 |
| Neuroticism | (Nagel et al., 2018) | Y | 449,484 |  |
| **Physical health** |  |  |  |  |
| BMI | (Yengo et al., 2018) | Y | 681,275 |  |
| Triglycerides | (Sinnott-Armstrong et al., 2021) |  | 363,228 |  |
| Type-2 Diabetes | (Suzuki et al., 2024) |  | 428,452 | 2,107,149 |
| IL-6 | (Sun et al., 2023) | NA | 54,219 |  |
| Insulin Resistance | (Oliveri et al., 2024) |  | 402,398 |  |
| Migraine | (Hautakangas et al., 2022) | NA | 102,084 | 771,257 |
| CRP | (Said et al., 2022) |  | 575,531 |  |
| Fasting Insulin | (Dupuis et al., 2010) |  | 38,238 |  |
| IL-1B | (Sun et al., 2023) |  | 54,219 |  |
| CAD | (Aragam et al., 2022) |  | 181,522 | 1,165,690 |
| Fasting Glucose | (Lagou et al., 2023) |  | 476,326 |  |
| IL-10 | (Sun et al., 2023) | NA | 54,219 |  |
| IL-12A | (Sun et al., 2023) | NA | 54,219 |  |
| HbA1c | (Sinnott-Armstrong et al., 2021) |  | 345,814 |  |
| TNF-alpha | (Sun et al., 2023) | NA | 54,219 |  |
| HDL | (Sinnott-Armstrong et al., 2021) |  | 332,323 |  |
| **Sleep and circadian-related** | | | | |
| Sleep midpoint | (Jones et al., 2019) | NA | 697,828 |  |
| Insomnia | (Jansen et al., 2019) |  | 1,331,010 |  |
| Relative amplitude | (Ferguson et al., 2018) | NA | 71,500 |  |
| Sleep duration | (Jansen et al., 2019) |  | 1,331,010 |  |
| Chronotype | (Jones et al., 2019) | NA | 697,828 |  |

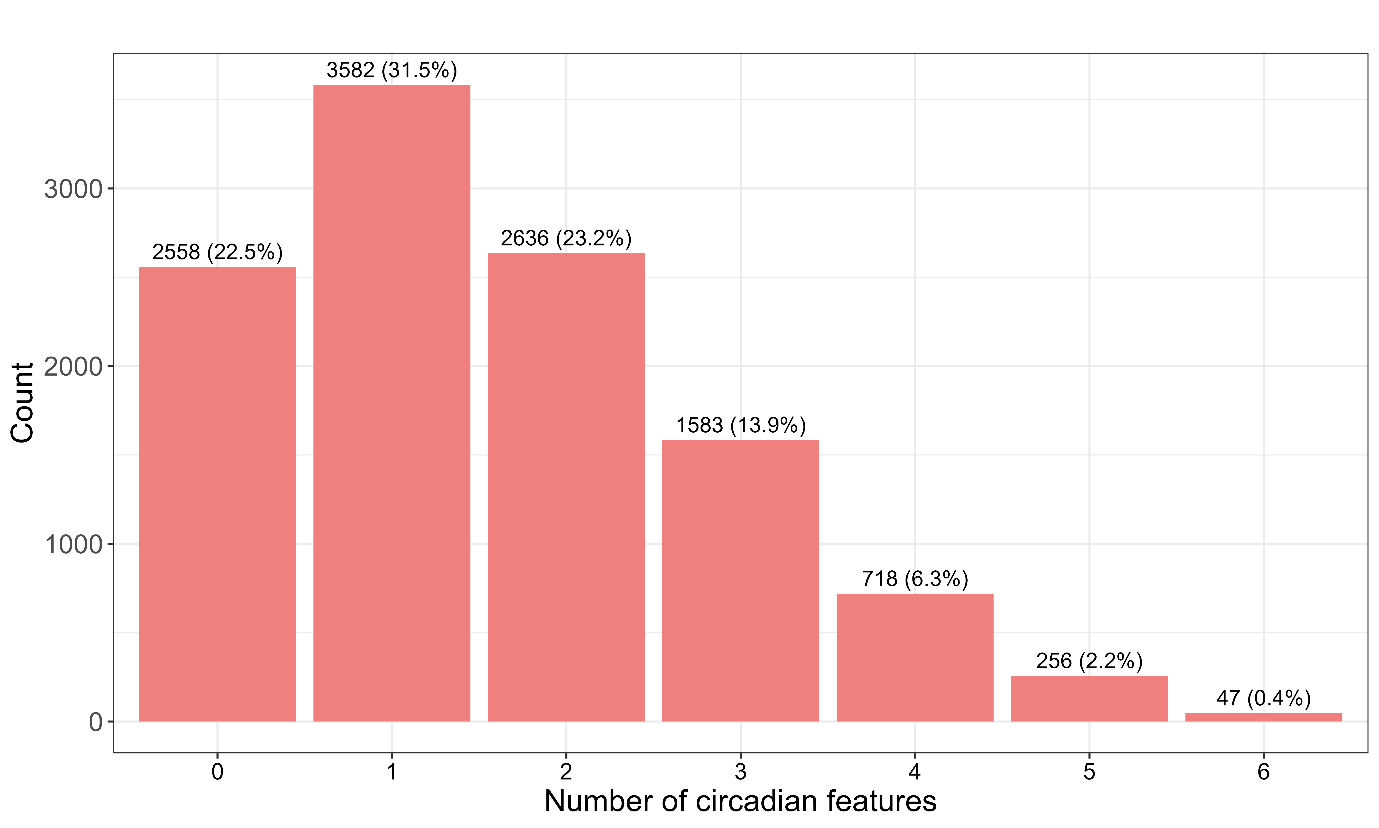

**Figure S2 Number of circadian criteria met in the whole sample (N=11,380).**

| **Table S3 Associations between the circadian subtype and polygenic risk scores and reported antidepressants side effects (n=2,604 with circadian cases; n=8,776 with non-circadian cases).** | | | | | |
| --- | --- | --- | --- | --- | --- |
|  | **OR** | **95%CI** | | ***p-value*** | ***p*Bonf** |
| *Mental disorders* |  |  |  |  |  |
| **ADHD** | **1.113** | **[1.064,** | **1.165]** | **0.000** | **Bonferroni corrected** |
| **Depression** | **1.113** | **[1.064,** | **1.164]** | **0.000** | **Bonferroni corrected** |
| **Bipolar** | **1.090** | **[1.042,** | **1.141]** | **0.000** | **Bonferroni corrected** |
| Schizophrenia | 1.073 | [1.020, | 1.128] | 0.006 | p < 0.05 |
| Neuroticism | 0.982 | [0.938, | 1.027] | 0.419 | Not significant |
| Autism | 1.038 | [0.992, | 1.087] | 0.105 | Not significant |
| *Physical health* |  |  |  |  |  |
| **BMI** | **1.093** | **[1.045,** | **1.144]** | **0.000** | **Bonferroni corrected** |
| CRP | 1.063 | [1.016, | 1.112] | 0.008 | p < 0.05 |
| **Triglycerides** | **1.104** | **[1.055,** | **1.155]** | **0.000** | **Bonferroni corrected** |
| **IL-6** | **1.075** | **[1.028,** | **1.125]** | **0.001** | **Bonferroni corrected** |
| HbA1c | 0.997 | [0.953, | 1.043] | 0.899 | Not significant |
| Fasting Insulin | 1.055 | [1.008, | 1.105] | 0.023 | p < 0.05 |
| IL-10 | 0.995 | [0.951, | 1.040] | 0.815 | Not significant |
| Migraine | 1.073 | [1.026, | 1.122] | 0.002 | p < 0.05 |
| TNFa | 0.991 | [0.948, | 1.036] | 0.703 | Not significant |
| IL-12a | 1.008 | [0.964, | 1.055] | 0.712 | Not significant |
| IL-1b | 1.033 | [0.987, | 1.080] | 0.159 | Not significant |
| Fasting Glucose | 1.018 | [0.973, | 1.066] | 0.439 | Not significant |
| HDL | 0.934 | [0.893, | 0.977] | 0.003 | p < 0.05 |
| CAD | 1.047 | [1.000, | 1.095] | 0.048 | p < 0.05 |
| T2D | 1.073 | [1.021, | 1.129] | 0.006 | p < 0.05 |
| **Insulin Resistance** | **1.084** | **[1.037,** | **1.134]** | **0.000** | **Bonferroni corrected** |
| *Sleep and circadian* |  |  |  |  |  |
| **Chronotype** | **0.679** | **[0.647,** | **0.712]** | **0.000** | **Bonferroni corrected** |
| **Sleep Midpoint** | **1.154** | **[1.103,** | **1.208]** | **0.000** | **Bonferroni corrected** |
| **Insomnia** | **1.076** | **[1.028,** | **1.126]** | **0.002** | **Bonferroni corrected** |
| Sleep Duration | 0.951 | [0.909, | 0.995] | 0.029 | p < 0.05 |
| Relative Amplitude | 1.062 | [1.016, | 1.111] | 0.008 | p < 0.05 |
| *Treatment response* |  |  |  |  |  |
| **SSRI efficacy** | **0.876** | **[0.821,** | **0.936]** | 0.000 | **Bonferroni corrected** |
| **SNRI efficacy** | **0.894** | **[0.826,** | **0.968]** | **0.006** | **Bonferroni corrected** |
| TCA efficacy | 0.945 | [0.836, | 1.067] | 0.362 | Not significant |
| *Notes.* For PGS analyses, covariates included age, sex, weight gain during episodes, and the 4 first PCs for population ancestry. For treatment response analyses, covariates included age and sex. ADHD: Attention-deficit Hyperactivity disorder; BMI: body mass index; T2D: Type-2 Diabetes; IL-6: Interleukin-6; CRP: C-reactive protein; IL-1b: Interleukin-1b; CAD: Coronary artery disease; IL-10: Interleukin-10; IL-12a: Interleukin-12a; TNF-a: Tumor Necrosis Factor-alpha; HDL: High-density lipoprotein. | | | | | |

| **Table S4 Sensitivity analyses, correcting for weight gain effect** | | | | | |
| --- | --- | --- | --- | --- | --- |
|  | **OR** | **95%CI** | | ***p-value*** | ***p*Bonf** |
| *Mental disorders* |  |  |  |  |  |
| **ADHD** | **1.086** | **[1.036,** | **1.138]** | **0.001** | **Bonferroni corrected** |
| **Depression** | **1.092** | **[1.043,** | **1.144]** | **0.000** | **Bonferroni corrected** |
| **Bipolar** | **1.083** | **[1.033,** | **1.134]** | **0.001** | **Bonferroni corrected** |
| Schizophrenia | 1.078 | [1.024, | 1.135] | 0.004 | p < 0.05 |
| Neuroticism | 0.991 | [0.946, | 1.038] | 0.697 | Not significant |
| Autism | 1.036 | [0.989, | 1.085] | 0.140 | Not significant |
| *Physical health* |  |  |  |  |  |
| BMI | 1.054 | [1.006, | 1.105] | 0.027 | p < 0.05 |
| CRP | 1.045 | [0.998, | 1.094] | 0.063 | Not significant |
| **Triglycerides** | **1.098** | **[1.048,** | **1.150]** | **0.000** | **Bonferroni corrected** |
| IL-6 | 1.075 | [1.027, | 1.126] | 0.002 | p < 0.05 |
| HbA1c | 1.002 | [0.957, | 1.050] | 0.917 | Not significant |
| Fasting Insulin | 1.048 | [1.000, | 1.099] | 0.050 | p < 0.05 |
| Interleukin_10 | 0.986 | [0.942, | 1.032] | 0.555 | Not significant |
| Migraine | 1.071 | [1.023, | 1.122] | 0.003 | p < 0.05 |
| TNF-a | 0.990 | [0.946, | 1.037] | 0.678 | Not significant |
| IL-12a | 1.011 | [0.966, | 1.059] | 0.640 | Not significant |
| IL-1b | 1.041 | [0.995, | 1.090] | 0.082 | Not significant |
| Fasting Glucose | 1.016 | [0.970, | 1.065] | 0.493 | Not significant |
| HDL | 0.945 | [0.903, | 0.990] | 0.016 | p < 0.05 |
| CAD | 1.047 | [0.999, | 1.096] | 0.054 | Not significant |
| T2D | 1.054 | [1.000, | 1.110] | 0.048 | p < 0.05 |
| Insulin Resistance | 1.068 | [1.020, | 1.118] | 0.005 | p < 0.05 |
| *Sleep and circadian PGS* |  |  |  |  |  |
| **Chronotype** | **0.675** | **[0.643,** | **0.710]** | **0.000** | **Bonferroni corrected** |
| **Sleep Midpoint** | **1.158** | **[1.106,** | **1.214]** | **0.000** | **Bonferroni corrected** |
| Insomnia | 1.065 | [1.017, | 1.116] | 0.008 | p < 0.05 |
| Sleep Duration | 0.956 | [0.913, | 1.001] | 0.054 | Not significant |
| Relative Amplitude | 1.060 | [1.012, | 1.110] | 0.013 | p < 0.05 |
| *Treatment response* |  |  |  |  |  |
| **SSRI efficacy** | **0.898** | **[0.840,** | **0.960]** | **0.002** | **Bonferroni corrected** |
| **SNRI efficacy** | **0.905** | **[0.835,** | **0.981]** | **0.015** | **Bonferroni corrected** |
| TCA efficacy | 0.943 | [0.833, | 1.067] | 0.353 | Not significant |
| *Notes.* For PGS analyses, covariates included age, sex, weight gain during episodes, and the 4 first PCs for population ancestry. For treatment response analyses, covariates included age and sex. ADHD: Attention-deficit Hyperactivity disorder; BMI: body mass index; T2D: Type-2 Diabetes; IL-6: Interleukin-6; CRP: C-reactive protein; IL-1b: Interleukin-1b; CAD: Coronary artery disease; IL-10: Interleukin-10; IL-12a: Interleukin-12a; TNF-a: Tumor Necrosis Factor-alpha; HDL: High-density lipoprotein. | | | | | |

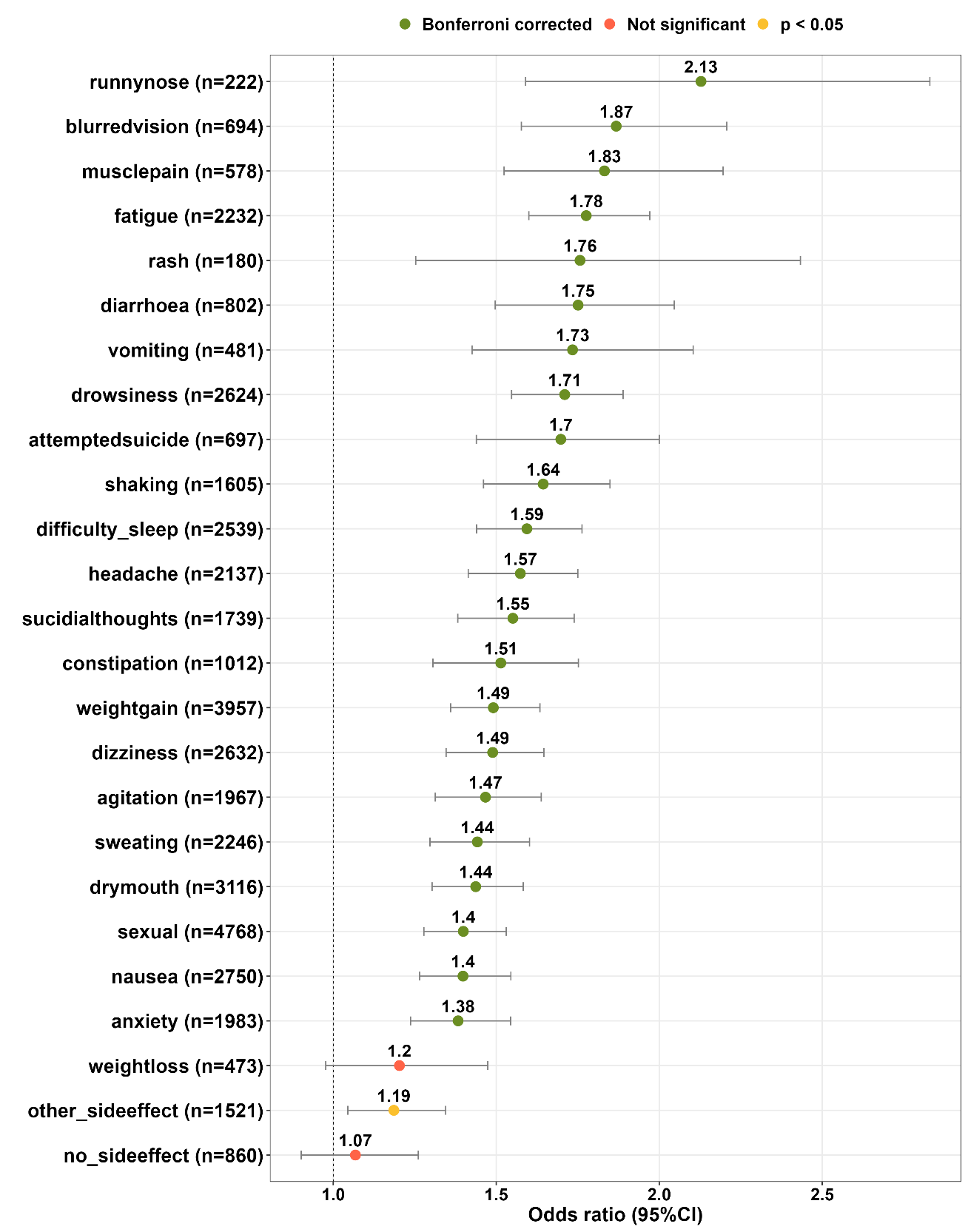

**Figure S3. Associations between the circadian subtype and self-reported side effects of medications (n=2,604 with circadian cases; n=8,776 with non-circadian cases).**

Notes: Results shown are separate logistic regression models for each variable with the circadian subtype, with covariates of age and sex. Significance levels: *Green*, Bonferroni-corrected (p<0.002 [0.05/25]); *Red*, Not significant. SNRI: Serotonin-norepinephrine reuptake inhibitor; SSRI: Selective serotonin reuptake inhibitors; TCA: Tricyclic antidepressants. Error bars represent 95% confidence interval.

| **Table S5 Associations between the circadian subtype and reported antidepressant side effects (n=2,604 with circadian cases; n=8,776 with non-circadian cases).** | | | | | |
| --- | --- | --- | --- | --- | --- |
|  | **OR** | **95% CI** | | ***p-value*** | ***p*Bonf** |
| **dry mouth (n=3116)** | **1.437** | **[1.303,** | **1.583]** | **0.000** | **Bonferroni corrected** |
| **sweating (n=2246)** | **1.442** | **[1.297,** | **1.602]** | **0.000** | **Bonferroni corrected** |
| **nausea (n=2750)** | **1.398** | **[1.265,** | **1.545]** | **0.000** | **Bonferroni corrected** |
| **vomiting (n=481)** | **1.734** | **[1.426,** | **2.104]** | **0.000** | **Bonferroni corrected** |
| **diarrhoea (n=802)** | **1.751** | **[1.497,** | **2.046]** | **0.000** | **Bonferroni corrected** |
| **constipation (n=1012)** | **1.514** | **[1.306,** | **1.752]** | **0.000** | **Bonferroni corrected** |
| **headache (n=2137)** | **1.574** | **[1.414,** | **1.750]** | **0.000** | **Bonferroni corrected** |
| **dizziness (n=2632)** | **1.489** | **[1.346,** | **1.646]** | **0.000** | **Bonferroni corrected** |
| **shaking (n=1605)** | **1.644** | **[1.461,** | **1.849]** | **0.000** | **Bonferroni corrected** |
| **muscle pain (n=578)** | **1.832** | **[1.524,** | **2.196]** | **0.000** | **Bonferroni corrected** |
| **drowsiness (n=2624)** | **1.710** | **[1.547,** | **1.889]** | **0.000** | **Bonferroni corrected** |
| **difficulty sleep (n=2539)** | **1.594** | **[1.440,** | **1.763]** | **0.000** | **Bonferroni corrected** |
| **anxiety (n=1983)** | **1.383** | **[1.237,** | **1.544]** | **0.000** | **Bonferroni corrected** |
| **agitation (n=1967)** | **1.467** | **[1.313,** | **1.638]** | **0.000** | **Bonferroni corrected** |
| **fatigue (n=2232)** | **1.776** | **[1.600,** | **1.971]** | **0.000** | **Bonferroni corrected** |
| **weight gain (n=3957)** | **1.491** | **[1.360,** | **1.634]** | **0.000** | **Bonferroni corrected** |
| **rash (n=180)** | **1.757** | **[1.253,** | **2.433]** | **0.001** | **Bonferroni corrected** |
| **weight loss (n=473)** | 1.203 | [0.977, | 1.474] | 0.078 | Not significant |
| **runny nose (n=222)** | **2.128** | **[1.590,** | **2.830]** | **0.000** | **Bonferroni corrected** |
| **sexual (n=4768)** | **1.399** | **[1.278,** | **1.531]** | **0.000** | **Bonferroni corrected** |
| **blurred vision (n=694)** | **1.868** | **[1.577,** | **2.207]** | **0.000** | **Bonferroni corrected** |
| **suicidal thoughts (n=1739)** | **1.551** | **[1.382,** | **1.739]** | **0.000** | **Bonferroni corrected** |
| **attempted suicide (n=697)** | **1.698** | **[1.439,** | **2.000]** | **0.000** | **Bonferroni corrected** |
| **other side effect (n=1521)** | 1.186 | [1.045, | 1.345] | 0.008 | p < 0.05 |
| **no side effect (n=860)** | 1.068 | [0.902, | 1.261] | 0.439 | Not significant |
| *Notes.* OR: odds ratio; CI: confidence interval | | | | | |

**Within-group comparison of antidepressant efficacy**

Of the 10 most common antidepressants, the circadian subtype group had tried significantly more antidepressants compared to the non-circadian subtype group (Mean=2.62±1.85 vs. Mean=2.22±1.68, respectively; *p*<0.001).

A linear mixed-effects model was conducted to compare efficacy ratings across antidepressant classes (SSRI, SNRI, TCA) within the circadian group and the non-circadian group separately, including a random intercept for participant ID. Pairwise contrasts showed no significant difference between SNRIs (*Mean=*2.10) and SSRIs (*Mean=* 2.15) (*estimate*=–0.05, *SE*=0.03, *z*=–1.98, *p* =0.143). In contrast, both SNRIs and SSRIs were rated as significantly more efficacious than TCAs (*Mean=*1.75). Specifically, SNRIs exceeded TCAs by 0.36 points (*SE*=0.04, *z*=9.78, *p*<0.001), and SSRIs exceeded TCAs by 0.41 points (*SE*=0.03, *z*=11.88, *p*<0.001). These results indicate that, within the circadian group, SSRIs and SNRIs demonstrate comparable efficacy, while TCAs are rated significantly lower.

Within the non-circadian subtype group, results indicated that SSRIs (*Mean=*2.24) were rated slightly but significantly higher than SNRIs (*Mean=*2.18) (*estimate*=–0.05, *SE*=0.02, *z*=–3.16, *p*=0.005). Both SSRIs and SNRIs were again rated more efficacious than TCAs (*Mean*=1.84). SNRIs exceeded TCAs by 0.34 points (*SE*=0.02, *z*=16.10, *p*<0.001), and SSRIs exceeded TCAs by 0.38 points (*SE*=0.02, *z*=19.73, *p*<0.001).

| **Table S6 Comparison between atypical depression and the circadian subtype of depression in AGDS** | | |
| --- | --- | --- |
|  | **Circadian vs. non circadian** | **Atypical vs. non atypical** |
| *Mental disorders* | | |
| ADHD | **++** | **++** |
| Depression | **++** | **+** |
| Bipolar disorder | **++** | ns |
| Schizophrenia | **+** | ns |
| Autism | ns | ns |
| Neuroticism | ns | **+** |
| Alzheimer’s | not tested | ns |
| *Physical health* | | |
| BMI | **++** | **++** |
| Triglycerides | **++** | ns |
| Type-2 Diabetes | **+** | **++** |
| Interleukin-6 | **++** | ns |
| Insulin Resistance | **++** | not tested |
| Migraine | **+** | not tested |
| C-Reactive Protein | **+** | **++** |
| Fasting Insulin | **+** | ns |
| Interleukin-1b | ns | ns |
| Coronary Artery Disease | **+** | **+** |
| Fasting Glucose | ns | ns |
| Interleukin-10 | ns | ns |
| Interleukin-12a | ns | ns |
| HbA1c | ns | ns |
| TNF-alpha | ns | ns |
| HDL | **-** | **--** |
| HOMA-IR | not tested | **+** |
| *Sleep and circadian PGS* | | |
| Sleep midpoint | **++** | not tested |
| Insomnia | **++** | not tested |
| Relative amplitude | **+** | not tested |
| Sleep duration | **-** | not tested |
| Chronotype | **--** | not tested |
| *Antidepressant response* | | |
| SSRI Efficacy | ns | ns |
| SNRI Efficacy | **--** | **--** |
| TCA Efficacy | **--** | **--** |
| *Notes. ++* indicates a positive association with a Bonferroni-corrected significant p-value; + indicates a positive association with a nominally significant p-value; **--** indicates a negative association with a Bonferroni-corrected significant p-value; **-** indicates a negative association with a nominally significant p-value; ns indicates non-significance. | | |

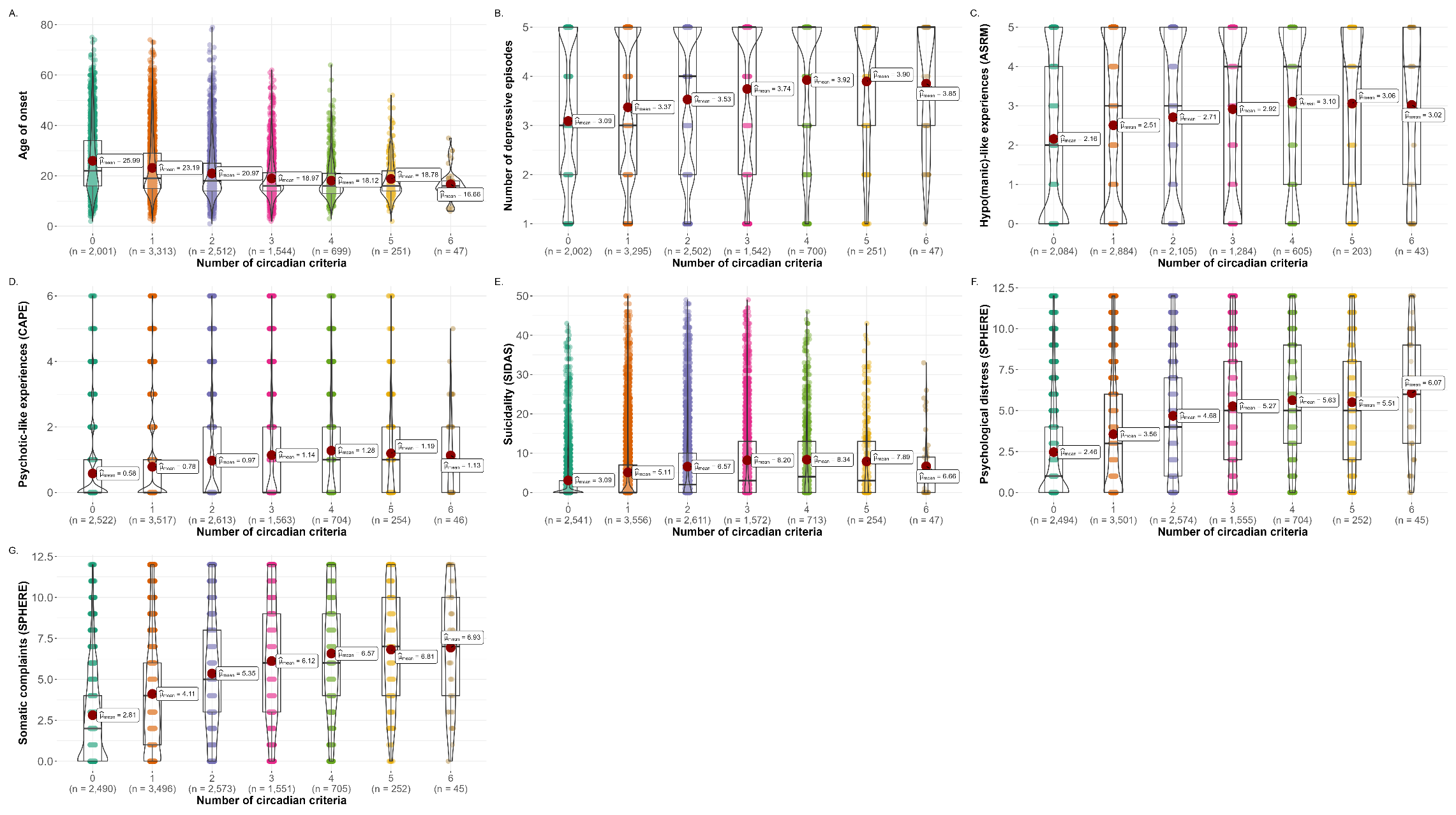

**Figure S4 Associations between number of criteria and clinical variables.**

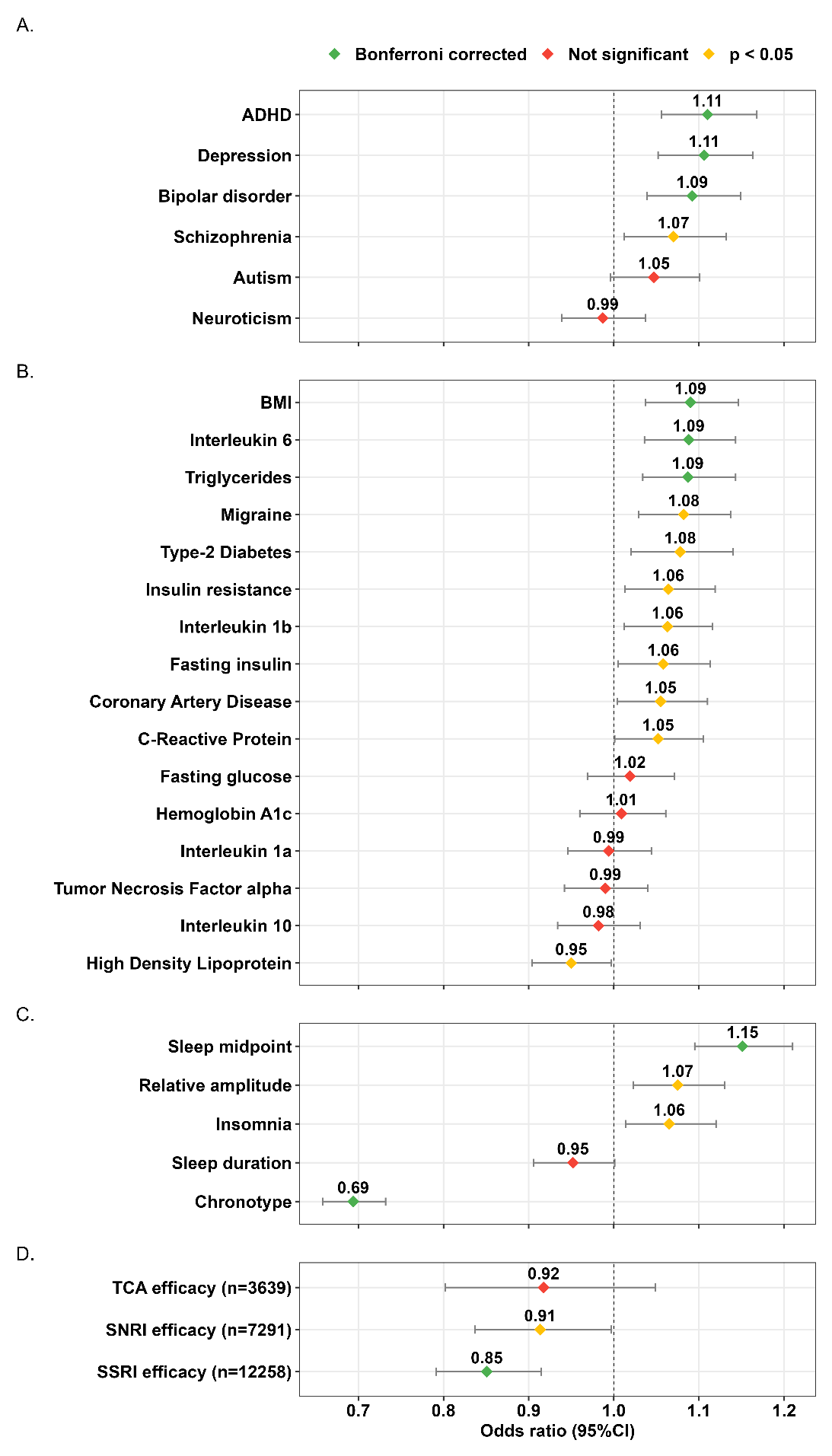
**Figure S5 Associations between the circadian subtype and polygenic risk scores for (A) mental disorders, (B) physical health, and (C) sleep and circadian factors and (D) treatment response restricted to those with data complete for all 6 circadian criteria (n=2,180 with circadian cases; n=6,548 with non-circadian cases).**

*Notes.* Results shown are separate logistic regression models for each variable with the circadian subtype, with covariates of age and sex for all models, and additionally the first 4 genetically inferred ancestry PCs for PGS models. Significance levels: *Green*, Bonferroni-corrected (at alpha level p<0.0019 [0.05/27] 6 mental disorder PGS, 16 physical health PGS, 5 sleep and circadian-related measures PGS), and at alpha level p<0.0167 [0.05/3] for treatment response); *Yellow*, Uncorrected (*p*<0.05); *Red*, Not significant. Error bars represent 95% confidence interval.

ADHD: Attention-Deficit Hyperactivity Disorder; BMI: Body Mass index; SNRI: Serotonin-norepinephrine reuptake inhibitor; SSRI: Selective serotonin reuptake inhibitors; TCA: Tricyclic antidepressants.

**Sensitivity analyses – restricted to those with complete data on all 6 circadian criteria**

To assess the robustness of these findings, we conducted sensitivity analyses restricting our sample size to those with complete data on all six circadian criteria (n=2,180 with circadian cases; n=6,548 with non-circadian cases). Similar patterns were observed with comparable effect sizes compared to the main analyses. Associations between the circadian subtype and mental health disorders PGS remained unchanged. Associations between the circadian subtype and physical health PGS remained unchanged, except for the association with insulin resistance PGS that did not survive Bonferroni correction (OR=1.06; 95% CI = [1.01, 1.12]; *p*=0.014) and that with Interleukin 1b PGS that reached nominal significance (OR=1.06; 95% CI = [1.01, 1.12]; *p*=0.015). Associations between the circadian subtype and sleep and circadian-related factors remained unchanged, except for the association with insomnia PGS that did not survive Bonferroni correction (OR=1.07; 95% CI = [1.01, 1.12]; *p*=0.013) and that with sleep duration PGS that did not reach significance (OR=0.95; 95% CI = [0.91, 1.00]; *p*=0.053). Association between the circadian subtype and self-reported SNRI efficacy did not survive Bonferroni correction (OR=0.91; 95% CI = [0.84, 1.00], *p*=0.042). Other associations with treatment response were unchanged from the main analyses.

| **Table S7 Associations between the circadian subtype and polygenic risk scores and reported antidepressants side effects restricted to those with data complete for all 6 circadian criteria (n=2,180 with circadian cases; n=6,548 with non-circadian cases).** | | | | | |
| --- | --- | --- | --- | --- | --- |
|  | **OR** | **95%CI** | | ***p-value*** | ***p*Bonf** |
| *Mental disorders* |  |  |  |  |  |
| **ADHD** | **1.110** | **[1.056,** | **1.168]** | **0.000** | **Bonferroni corrected** |
| **Depression** | **1.106** | **[1.052,** | **1.163]** | **0.000** | **Bonferroni corrected** |
| **Bipolar** | **1.092** | **[1.039,** | **1.149]** | **0.001** | **Bonferroni corrected** |
| Schizophrenia | 1.070 | [1.012, | 1.132] | 0.017 | p < 0.05 |
| Neuroticism | 0.987 | [0.939, | 1.037] | 0.602 | Not significant |
| Autism | 1.047 | [0.996, | 1.101] | 0.073 | Not significant |
| *Physical health* |  |  |  |  |  |
| **BMI** | **1.090** | **[1.037,** | **1.146]** | **0.001** | **Bonferroni corrected** |
| CRP | 1.052 | [1.001, | 1.105] | 0.046 | p < 0.05 |
| **Triglycerides** | **1.087** | **[1.043,** | **1.143]** | **0.001** | **Bonferroni corrected** |
| **IL-6** | **1.088** | **[1.036,** | **1.143]** | **0.001** | **Bonferroni corrected** |
| HbA1c | 1.009 | [0.960, | 1.061] | 0.725 | Not significant |
| Fasting Insulin | 1.058 | [1.005, | 1.113] | 0.030 | p < 0.05 |
| IL-10 | 0.982 | [0.934, | 1.031] | 0.462 | Not significant |
| Migraine | 1.082 | [1.029, | 1.137] | 0.002 | p < 0.05 |
| TNFa | 0.990 | [0.942, | 1.040] | 0.679 | Not significant |
| IL-12a | 0.994 | [0.946, | 1.044] | 0.811 | Not significant |
| IL-1b | 1.063 | [1.012, | 1.116] | 0.015 | p < 0.05 |
| Fasting Glucose | 1.019 | [0.969, | 1.071] | 0.474 | Not significant |
| HDL | 0.950 | [0.904, | 0.997] | 0.039 | p < 0.05 |
| CAD | 1.055 | [1.004, | 1.110] | 0.034 | p < 0.05 |
| T2D | 1.078 | [1.020, | 1.140] | 0.006 | p < 0.05 |
| Insulin Resistance | 1.064 | [1.013, | 1.119] | 0.014 | p < 0.05 |
| *Sleep and circadian* |  |  |  |  |  |
| **Chronotype** | **0.694** | **[0.658,** | **0.732]** | **0.000** | **Bonferroni corrected** |
| **Sleep Midpoint** | **1.151** | **[1.095,** | **1.210]** | **0.000** | **Bonferroni corrected** |
| Insomnia | **1.065** | **[1.014,** | **1.120]** | **0.013** | p < 0.05 |
| Sleep Duration | 0.952 | [0.906, | 1.001] | 0.053 | Not significant |
| Relative Amplitude | 1.075 | [1.023, | 1.130] | 0.004 | p < 0.05 |
| *Treatment response* |  |  |  |  |  |
| **SSRI efficacy** | **0.851** | **[0.792,** | **0.915]** | **0.000** | **Bonferroni corrected** |
| SNRI efficacy | 0.913 | [0.837, | 0.997] | 0.042 | p < 0.05 |
| TCA efficacy | 0.918 | [0.802, | 1.049] | 0.208 | Not significant |
| *Notes.* For PGS analyses, covariates included age, sex, weight gain during episodes, and the 4 first PCs for population ancestry. For treatment response analyses, covariates included age and sex. ADHD: Attention-deficit Hyperactivity disorder; BMI: body mass index; T2D: Type-2 Diabetes; IL-6: Interleukin-6; CRP: C-reactive protein; IL-1b: Interleukin-1b; CAD: Coronary artery disease; IL-10: Interleukin-10; IL-12a: Interleukin-12a; TNF-a: Tumor Necrosis Factor-alpha; HDL: High-density lipoprotein. | | | | | |

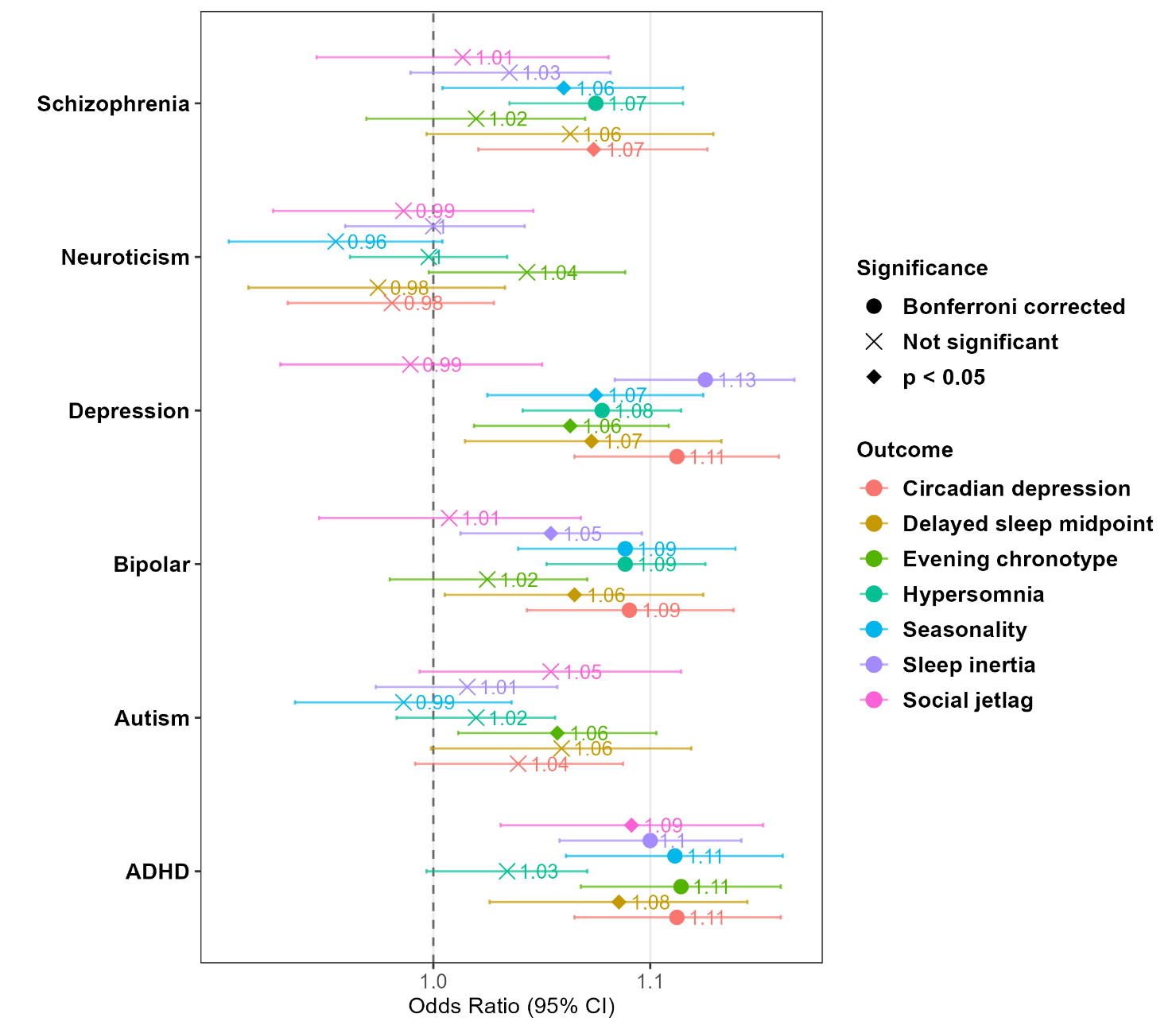
**Figure S6 Associations between each binary circadian criteria and polygenic risk scores for mental disorders.**

*Notes.* Results shown are separate logistic regression models for each variable with the circadian subtype, with covariates of age and sex for all models, and additionally the first 4 genetically inferred ancestry PCs for PGS models. Significance levels: Bonferroni-corrected (at alpha level p<0.0019 [0.05/27] 6 mental disorder PGS, 16 physical health PGS, 5 sleep and circadian-related measures PGS). Error bars represent 95% confidence interval. ADHD: Attention-Deficit Hyperactivity Disorder.

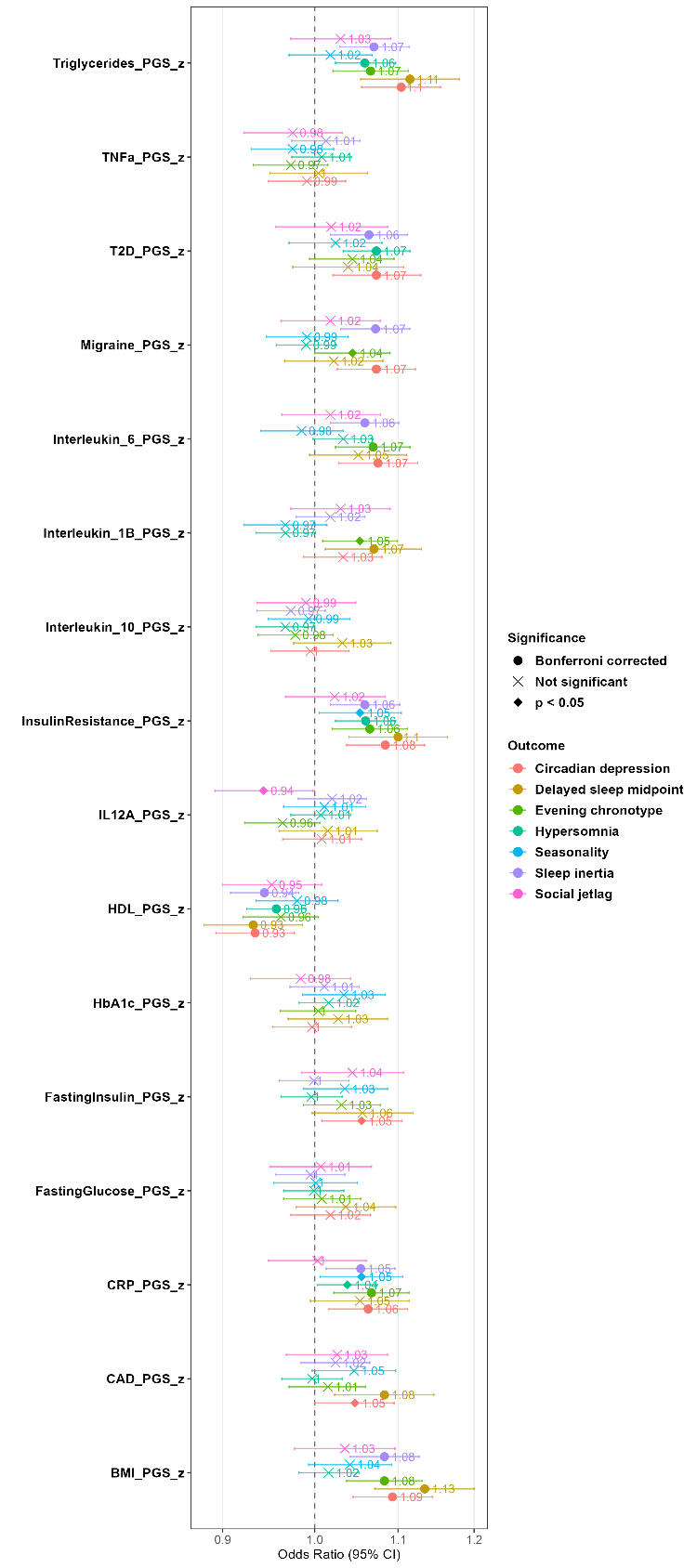
**Figure S7 Associations between each binary circadian criteria and polygenic risk scores for physical health.**

*Notes.* Results shown are separate logistic regression models for each variable with the circadian subtype, with covariates of age and sex for all models, and additionally the first 4 genetically inferred ancestry PCs for PGS models. Significance levels: Bonferroni-corrected (at alpha level p<0.0019 [0.05/27] 6 mental disorder PGS, 16 physical health PGS, 5 sleep and circadian-related measures PGS). Error bars represent 95% confidence interval.

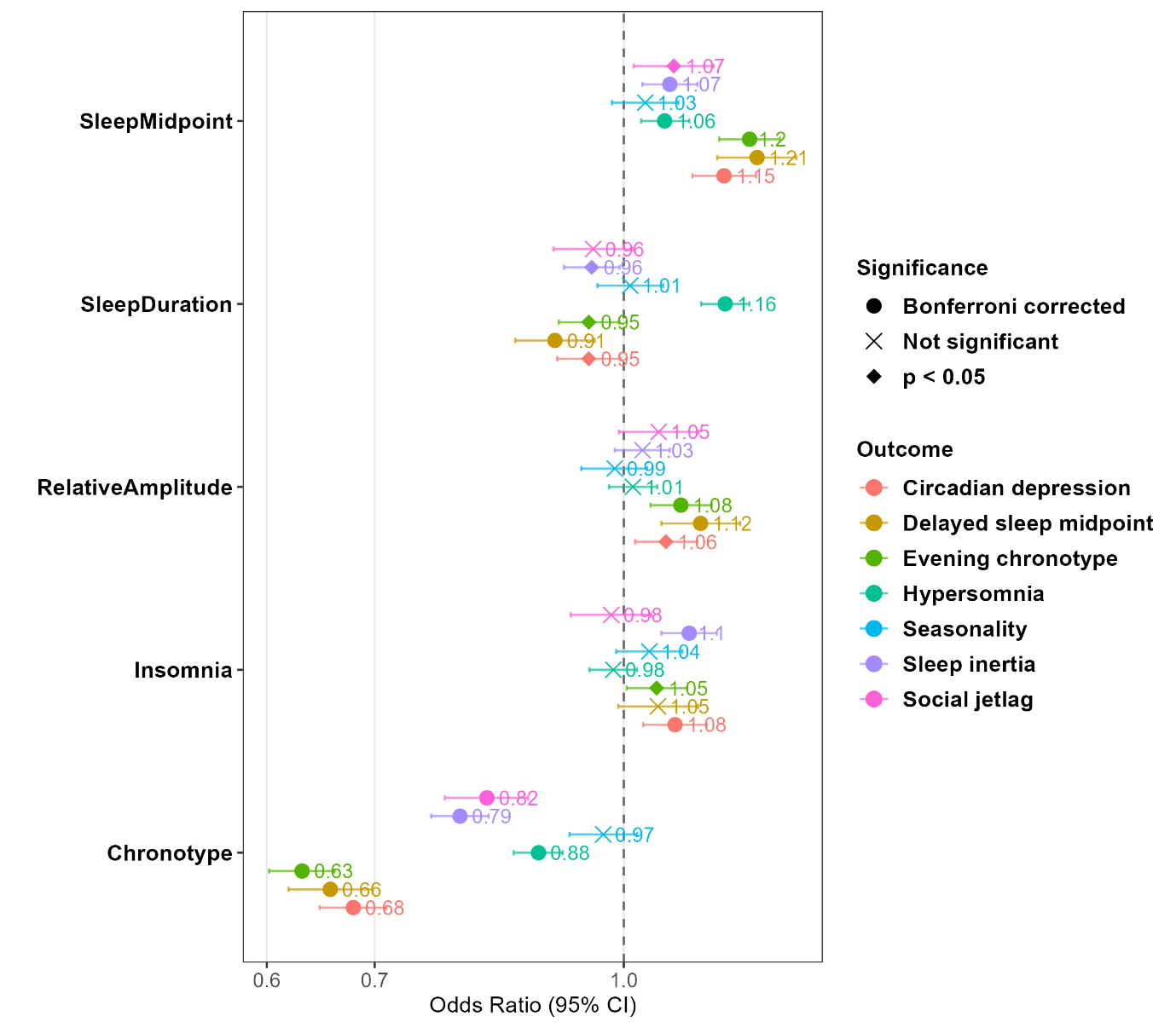
**Figure S8 Associations between each binary circadian criteria and polygenic risk scores for sleep and circadian factors.**

*Notes.* Results shown are separate logistic regression models for each variable with the circadian subtype, with covariates of age and sex for all models, and additionally the first 4 genetically inferred ancestry PCs for PGS models. Significance levels: Bonferroni-corrected (at alpha level p<0.0019 [0.05/27] 6 mental disorder PGS, 16 physical health PGS, 5 sleep and circadian-related measures PGS). Error bars represent 95% confidence interval.

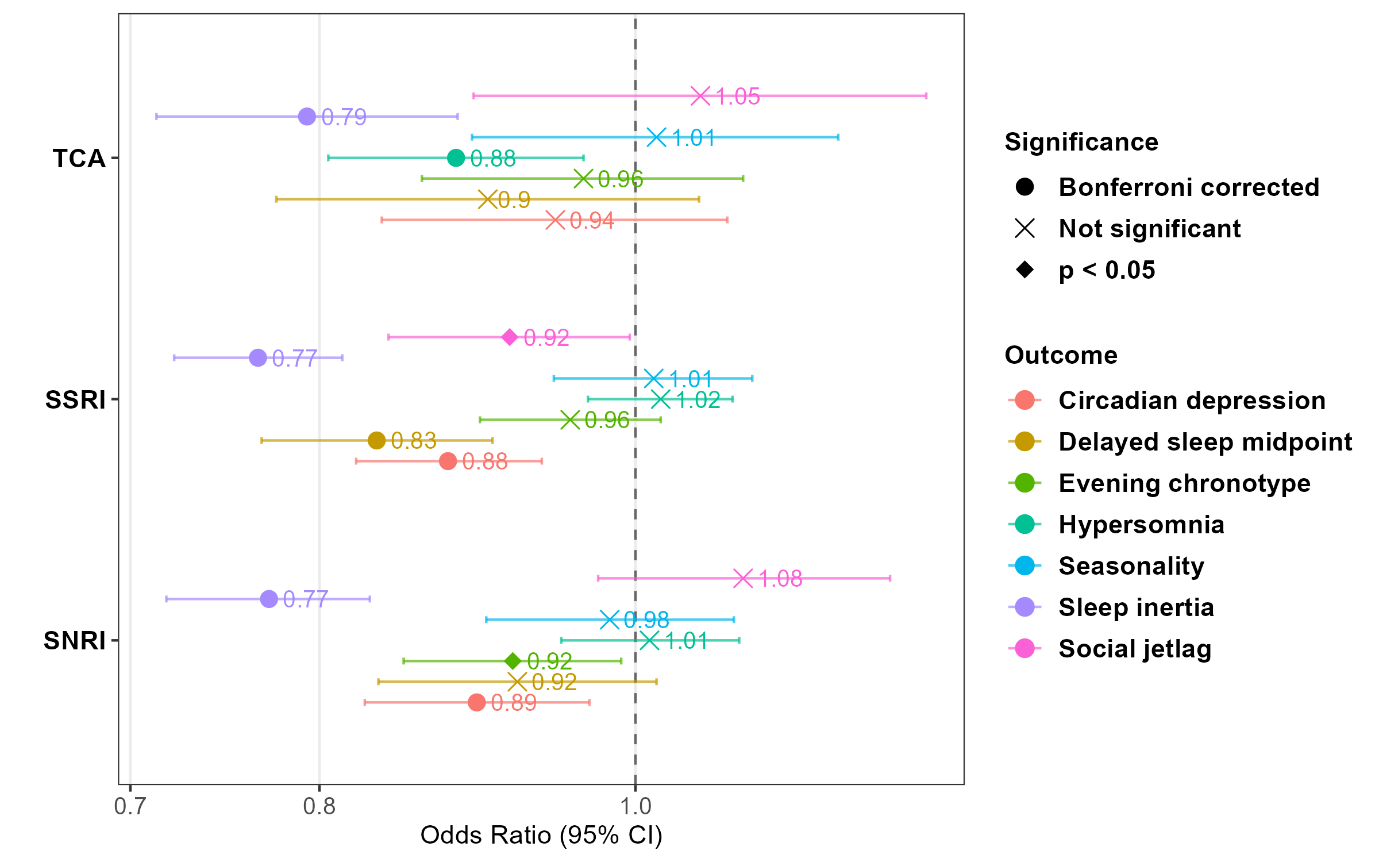
**Figure S9 Associations between each binary circadian criteria and self-reported antidepressant efficacy.**

*Notes.* Results shown are separate logistic regression models for each variable with the circadian subtype, with covariates of age and sex for all models, and additionally the first 4 genetically inferred ancestry PCs for PGS models. Significance levels: Bonferroni-corrected at alpha level p<0.0167 [0.05/3]. Error bars represent 95% confidence interval. SNRI: Serotonin-norepinephrine reuptake inhibitor; SSRI: Selective serotonin reuptake inhibitors; TCA: Tricyclic antidepressants.
